# Epidemiology, temporal trends, and fibrosis risk stratification in metabolic dysfunction-associated steatotic liver disease in UK primary care: a population-based cohort and nested case-control study

**DOI:** 10.64898/2026.07.15.26358136

**Authors:** Huei-Tyng Huang, Michael Hewitt, Wenhao Li, Laura Temperley, Naveed Sattar, William Alazawi

**Author notes:** (corresponding author) **Address of Correspondence:** Professor William Alazawi, 4 Newark Street, Blizard Institute, Queen Mary University of London, E1 2AT, United Kingdom.

## Abstract

**Background:** We sought to determine the changing prevalence, incidence, and temporal trends in real-world, recorded diagnoses of metabolic dysfunction-associated steatotic liver disease (MASLD) and assess availability of fibrosis risk stratification following awareness campaigns and guideline updates over the last decade.

**Methods:** This population-based cohort study identified MASLD diagnoses made between 2003-2022 in the UK primary-care Clinical Practice Research Datalink (CPRD) to estimate prevalence and incidence. A nested case-control analysis, utilising 1:4 age-, sex-, and general practice-matched controls, assessed clinical characteristics, availability of Fibrosis-4 (Fib-4) components, and its temporal trend pre- and post-2015.

**Findings:** 11.7 million individuals were active in CPRD in 2022. 365,797 comprised the study cohort of people with a MASLD diagnosis (matched to 1,460,288 controls). From 2012-2022, recorded MASLD prevalence rose from 0.52% [N=51,028] to 2.42% [N=283,762] (p<0.001); recorded incidence doubled from 1.60 to 3.31 per 1000 person-years (p<0.001). People with MASLD diagnosis had a higher prevalence of type 2 diabetes (21.0% [N=76,640] vs 7.7% [N=112,812]) and hypertension (35.3% [N=129,156] vs 18.7% [N=273,502]). People of South Asian ethnicity were overrepresented in MASLD cohort but had the lowest availability of Fib-4 components (14.6% [N=4,467]; adjusted odds ratio 0.67, 95% CI: 0.65–0.70, vs White). Overall, Fib-4 availability increased pre- to post-2015 (4.3% [N=4,945] to 22.8% [N=56,634]). Among those with a calculable score, fewer South Asian individuals had indeterminate/high risk (18.6% [N=833] vs 35.3% [N=15,115] in White individuals, p<0.001).

**Interpretation:** Recorded MASLD prevalence has increased 5-fold in a decade, yet a diagnostic gap persists. Fibrosis risk stratification has improved, but remains low and is potentially inequitable for people of South Asian ethnicity.

**Funding:** Merck Sharp & Dohme LLC, a subsidiary of Merck & Co., Inc., Rahway, NJ, USA; Barts Charity.

## Introduction

Metabolic dysfunction-associated steatotic liver disease (MASLD) is the most common cause of chronic liver disease worldwide^1^ and is closely associated with metabolic co-morbidities such as type 2 diabetes mellitus (T2DM) and obesity. The current estimated global prevalence of MASLD is 30% (95% confidence intervals CI: 28%–32%)^1^ based on meta-analysis of consistent findings from carefully-selected cohort studies of varying size, though substantial geographical data gaps remain. However, it is widely recognised that there is a significant diagnostic gap with under-diagnosis in health record data when compared to cohort studies. This means that patients with clinically-significant disease who may be amenable to intervention including with novel, recently-approved therapies, may not be linked to care providers.

MASLD comprises a wide spectrum of liver disease severity ranging from simple steatosis through to metabolic-dysfunction associated steatohepatitis (MASH), fibrosis, cirrhosis and hepatocellular carcinoma (HCC)^2^. The degree of liver fibrosis predicts liver-related outcomes and care pathways include early fibrosis risk stratification using scores such as the Fibrosis-4 (Fib-4), usually in primary care where most diagnoses are made. A large primary care real-world data (RWD) study reported an overall prevalence of diagnosed non-alcoholic fatty liver disease (NAFLD) in four European countries to be <3% in 2015 (0.6% in the United Kingdom, UK) with the data required to calculate Fib-4 available in less than half of people with the diagnosis (11.3% in the UK)^1^.

The last ten years have seen much activity to raise awareness and understanding of MASLD reflected in guidance and recommendations^3^. It is therefore reasonable to hypothesise that this activity should have led to a rise in prevalence of recorded MASLD diagnosis to a level closer to that estimated by cohort studies and that clinicians are performing more risk-stratification in the form of Fib-4. In this study, we used the Clinical Practice Research Datalink (CPRD), a comprehensive repository of primary care records for over 19 million currently registered individuals, more than a quarter of the UK population^4^, to test our hypothesis.

Large-scale RWD derived from routine clinical practice offer a powerful tool to study common diseases in the general population^5,6^ especially in territories where primary care is a gateway into clinical services and does not require co-payment for access. While data veracity and completeness cannot be assured, RWD can balance potential limitations of carefully selected cohort studies that may lack ethnic or socio-economic diversity^7^. CPRD contains routinely collected anonymised electronic health records (EHRs) from over 2,000 general practices (GP) across the UK (a country with a high obesity prevalence), capturing demographic, clinical, and laboratory data for millions of people with long follow-up. For example, the sex and age distribution match national demographics: 50.1% female, 20.0% aged <18, 62.6% aged 18–64, and 17.3% aged ≥65^8^. Within the limitation of RWD, CPRD can be used to estimate prevalence and incidence of recorded diagnoses, temporal trends, and insight into clinical practices^8^.

The objectives of the current study were to: 1) determine the prevalence and incidence of diagnosed MASLD in a large, unselected population; 2) characterise temporal trends between 2003 and 2022; and 3) investigate whether the data required to calculate the Fib-4 score after 2015 are more available in GP than before 2015.

## Methods

### Study Design

A dual analytical design was used to address distinct epidemiological questions. A retrospective population-based cohort design was utilised to estimate the annual prevalence and incidence of MASLD among all active adult individuals in the CPRD. A nested case-control design was utilised to investigate clinical characteristics and evaluate temporal and ethnicity-related trends in fibrosis risk stratification.

### Setting

This study is based on data from the CPRD obtained under licence from the UK Medicines and Healthcare products Regulatory Agency (MHRA). The data is provided by patients and collected by the National Health Service (NHS) as part of their care and support. The unique study protocol with ethical approval (ID: 24_004153) was approved by the CPRD under the Multi Study License (MSL) at Queen Mary University of London (QMUL). Data extraction was stored within the secure Data Safe Haven (DSH) at QMUL.

### Participants

Individuals who entered the CPRD Aurum database between 01/01/2003 and 31/12/2022 were included in the prevalence and incidence estimates, using the most recent anonymised data release version at the time of data extraction (December 2024). Study size was determined by the total number of eligible individuals registered within the database during the study period. Those with less than one year follow-up after CPRD registration were excluded. Patients aged 18 and above at the time of MASLD diagnosis were included, according to diagnostic codes and the date of diagnosis. Those with the following liver-related diagnoses made before MASLD diagnosis were excluded: haemochromatosis, autoimmune liver disease, chronic viral hepatitis, primary biliary cholangitis, primary sclerosing cholangitis, Wilson’s disease, and alpha-1 antitrypsin deficiency. Each person with MASLD was randomly matched with 1:4 ‘non-exposed’ control patients who did not have a MASLD diagnosis, based on sex, age (±5 years), and registered primary care practice. The study flow chart is summarised in **Supplementary Figure 1**.

### Variables and measurement

NAFLD is the most practical coding proxy for MASLD in EHRs^9^, and published code lists were used to curate the diagnosis in this study^10^. This approach is supported by recent studies demonstrating high concordance (all >94%) between NAFLD coding and MASLD diagnosis in multiple studies^11,12^. Medical codes for inclusion and exclusion criteria were referenced from Health Data Research UK (HDRUK) Phenotype library (https://phenotypes.healthdatagateway.org/) and the CPRD in-house code browser (**Supplementary Table 1**).

We extracted the following clinical data if recorded between 2 years prior to and 6 months after index date: body mass index (BMI), alanine transaminase (ALT), aspartate transaminase (AST), platelet counts, gamma-glutamyl transferase (GGT), Haemoglobin A1c (HbA1c), cholesterol, triglycerides, low-density lipoprotein (LDL) levels, and cardiovascular risk score QRISK2. The Fib-4 score was calculated through the formula 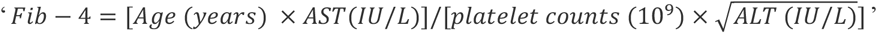 for those in whom the relevant blood results could be extracted^13^. The record closest to the index date was selected when multiple records were found. Patients’ self-reported ethnicity was used and categorised using codes. Ethnic groups were collapsed into five categories: White, Black, South Asian, Other Asian, and mixed/others, reflecting the ethnic composition of the UK. History of T2DM, hypertension, smoking, and excess alcohol use was defined as a record occurring any time before or at the index date. For the matched controls, we extracted the lab records closest to 2022 to reflect the most up-to-date GP landscape, and comorbidities were defined as having any CPRD records. Code lists were generated using the HDRUK Phenotype Library and CPRD code browser (**Supplementary Table 2**).

### Statistical methods

Point prevalence was estimated for December 31^st^ of each year. It was defined as the total number of recorded people with MASLD who were still active in CPRD at or before 31^st^ December of each calendar year, divided by the total number of active patients in the database. In the UK, primary care is a universal health system where nearly all residents register with a local GP. Consequently, in CPRD, ‘active’ refers to individuals who are registered with a contributing GP and therefore their records are available during the specified time frame, representing the general public. Incidence was defined as the number of individuals newly diagnosed with MASLD, divided by the number of person-years at risk. The annual incidence proportion was also estimated to account for potential follow-up bias among included populations. It was defined as the number of newly diagnosed individuals with MASLD in a calendar year divided by the individuals at risk during the same year. Prevalence and the annual incidence proportion were fitted using logistic regressions, and the incidence rate was fitted using Poisson regression. Subgroups by sex and age categories (18–29, 30–39, 40–49, 50–59, 60–69, 70–79, ≥80) were also analysed. The age of individuals was computed on 1^st^ January of the year of interest.

Continuous variables were reported as medians with interquartile range (IQRs), and categorical variables were recorded as percentages. Differences in characteristics between MASLD and matched controls were tested by Wilcoxon’s rank-sum test for continuous variables and the chi-squared test for categorical variables. We selected 2015 as a temporal cut-point to align with the end date of a previous primary care analysis^10^, allowing for assessment of progress in the subsequent period in which there was by increasing awareness and recently updated clinical guidelines.

To evaluate factors associated with the availability of a calculable Fib-4 score, univariable and multivariable odds ratios (ORs) were calculated using logistic regression. To avoid overadjustment, multivariable models were specifically constructed to assess the effect of ethnicity on Fib-4 availability. Confounders were defined a priori based on clinical knowledge; therefore, the model assessing the exposure of ethnicity was adjusted for age and sex, intentionally excluding comorbidities to avoid adjusting for metabolic mediators. A complete-case analysis was used, and a sensitivity analysis was performed by excluding individuals aged <35 to assess the robustness of risk stratification. All analyses were conducted using *RStudio (version 4*.*5*.*2)* provided in the DSH at QMUL.

### Declarations and ethics approval

The Methods are written following the Strengthening the reporting of observational studies in epidemiology (STROBE) guideline for observational studies^14^. The ethics were approved by the CPRD Research Data Governance committee, and a waiver of informed consent was granted because the data that CPRD receives from GPs is pseudonymised at source, and GPs do not need to seek individual patient consent when they share data with CPRD.

## Results

### Study population and characteristics

A total of 11,750,288 people were active in CPRD Aurum in 2022. 365,797 comprised the study cohort of people with MASLD diagnosis based on the inclusion and exclusion criteria and 1,460,288 matched controls, corresponding to a matching ratio of 3.99:1 (**Supplementary Figure 1**). The median age of people with MASLD diagnosis was 53 years (IQR: 43–63) and 51.8% (N=189,483) were male (**Table 1**). Compared with matched controls, people with MASLD diagnosis had a significantly higher median BMI (31.3 kg/m^2^, IQR: 27.9–35.6 vs 26.7 kg/m^2^, IQR: 23.5–30.7; p<0.001). 11.0% of people with MASLD diagnosis were of South Asian ethnicity; 1.5 times the proportion in matched controls (6.9% in the UK 2021 census^15^). 21.0% (N=76,640) of people with MASLD diagnosis had T2DM compared to 7.7% (N=112,812) of controls (p<0.001), and 35.3% (N=129,156) had hypertension compared to 18.7% (N=273,502) controls (p<0.001). Among those with T2DM, the mean HbA1c was 7.8% in MASLD and 7.5% in controls (p<0.001). Any smoking history was more common in people with MASLD diagnosis (54.4% [N=198,979] vs 40.7% [N=594,364]; p<0.001). While patients with coded alcohol excess or alcohol-related liver disease were excluded, 1.9% and 3.0% of people with MASLD had an ‘excess alcohol use’ record within 2 and 5 years respectively of the MASLD diagnosis. This low proportion of people with MASLD who also had records of excess alcohol use did not affect subsequent results (**Supplementary Table 3**). Liver-related laboratory measurements (the closest to index date within -2 years to +6 months of MASLD diagnosis) were higher in cases compared to controls (ALT, AST and GGT all p<0.001).

**Table 1.** Descriptive characteristics of MASLD patients and matched controls in the CPRD. For categorical variables, records were reported as counts (in percentages); for continuous variables, records were reported as medians (with IQRs).

|  | MASLD (N = 365,797) | Ctrl (N = 1,460,288) | Test of difference (p-value) |
| --- | --- | --- | --- |
| Age (years) | 53 (43 – 63) | 54 (40 – 65) | <0.001 |
| Sex – male | 51.8% | 52.6% | <0.001 |
| BMI (in kg/m<sup>2</sup>) | 31.3 (27.9 – 35.6) | 26.7 (23.5 – 30.7) | <0.001 |
| Ethnicity |  |  |  |
| White | 215,220 (77.9%) | 726,273 (82.2%) | <0.001 |
| Black | 11,469 (4.2%) | 44,322 (5.0%) |  |
| South Asian | 30,505 (11.0%) | 63,353 (7.2%) |  |
| Other Asian | 14,078 (5.1%) | 34,420 (3.9%) |  |
| Mixed / Others | 4,928 (1.7%) | 15,334 (1.7%) |  |
| N/A | 89,597 (-) | 576,586 (-) |  |
| History of comorbidities |  |  |  |
| Had T2DM | 76,640 (21.0%) | 112,812 (7.7%) | <0.001 |
| Had hypertension | 129,156 (35.3%) | 273,502 (18.7%) | <0.001 |
| Smoked | 198,979 (54.4%) | 594,364 (40.7%) | <0.001 |
| Had excess alcohol use | 17,493 (4.8%) | 37,330 (2.6%) | <0.001 |
| Lab measurements |  |  |  |
| ALT (IU/L) | 38 (24 – 62) | 21 (15 – 30) | <0.001 |
| AST (IU/L) | 32 (24 – 45) | 23 (19 – 29) | <0.001 |
| Platelets (10<sup>9</sup>/L) | 255 (214 – 302) | 256 (215 – 303) | <0.001 |
| GGT (IU/L) | 59 (34 – 115) | 29 (19 – 51) | <0.001 |
| HbA1c (%) | 5.6 (4.5 – 6.2) | 5.5 (5.1 – 5.9) | <0.001 |
| Cholesterol (mmol/L) | 5.0 (4.2 – 5.9) | 4.9 (4.2 – 5.7) | <0.001 |
| Triglycerides (mmol/L) | 1.8 (1.3 – 2.5) | 1.3 (1.0 – 1.9) | <0.001 |
| LDL (mmol/L) | 2.9 (2.2 – 3.6) | 2.9 (2.2 – 3.6) | 0.045 |
| **Fibrosis-4 score** | **61,579 (16.8%)** | **141,882 (9.7%)** | **<0.001** |
| Low risk (<1.30) | 41,222 (66.9%) | 76,672 (54.0%) | - |
| Indeterminate risk | 16,359 (26.6%) | 53,251 (36.4%) | - |
| High risk (>2.67) | 3,998 (6.5%) | 11,959 (8.2%) | - |
| **QRISK2 score** | **101,037 (27.6%)** | **209,965 (28.1%)** | **<0.001** |
| Low-risk (<10%) | 55,092 (54.5%) | 206,407 (50.3%) | - |
| Medium risk | 27,240 (27.0%) | 117,980 (28.8%) | - |
| High risk (>20%) | 18,705 (18.5%) | 85,578 (20.9%) | - |

### Prevalence of recorded MASLD

The recorded point prevalence of MASLD in the CPRD rose significantly and consistently over the study period, rising from 0.02% (N=1,300) in 2003 to 2.42% (N=283,762) in 2022 (overall trend: p<0.001) (**Figure 1**). This increasing trend was also observed across all age and sex subgroups (**Figure 2**). Prevalence rose with age, increasing steadily from the 18–29 age group and peaking in the 60–69 age group, across all time points.

**Figure 1.**
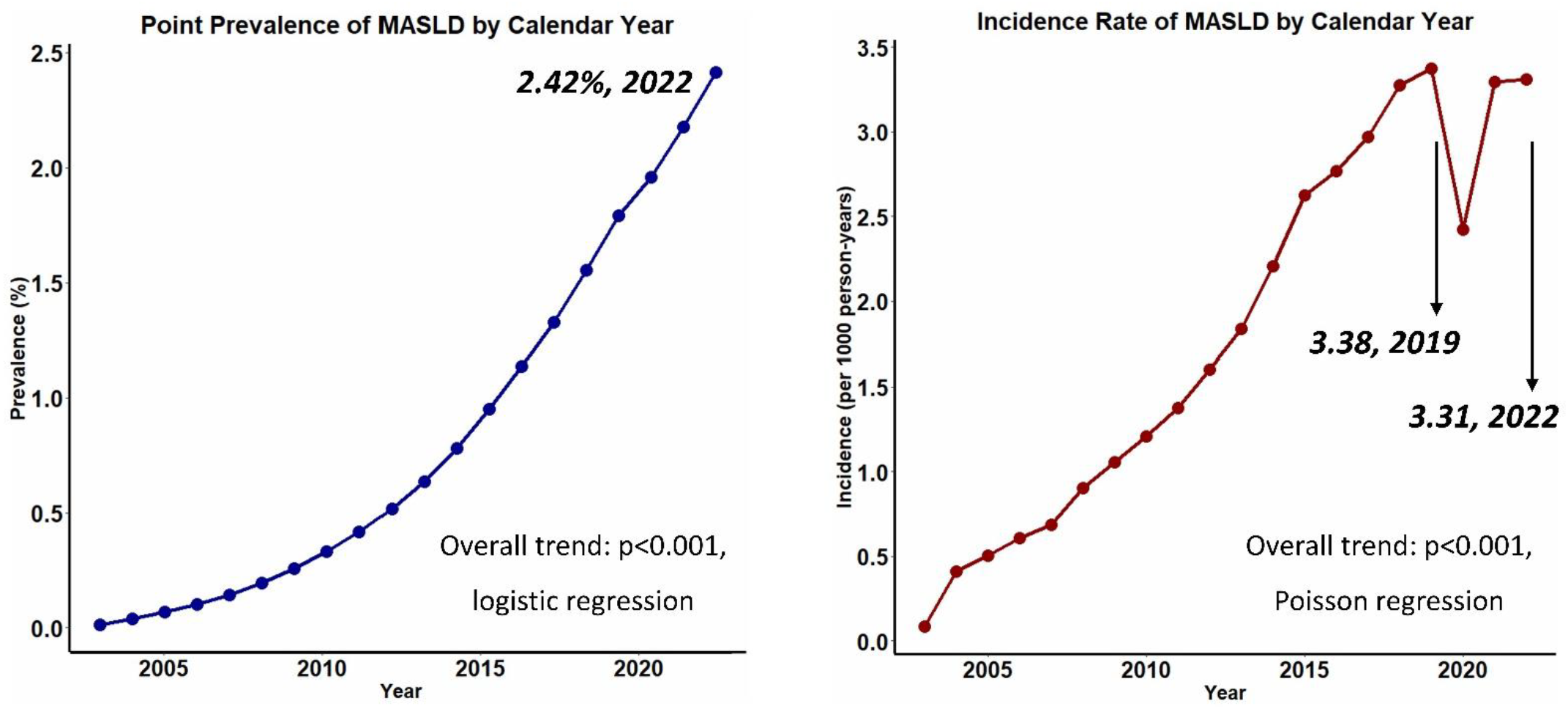
Recorded prevalence and incidence of MASLD in the CPRD. Left: Prevalence. Right: Incidence.

**Figure 2.**
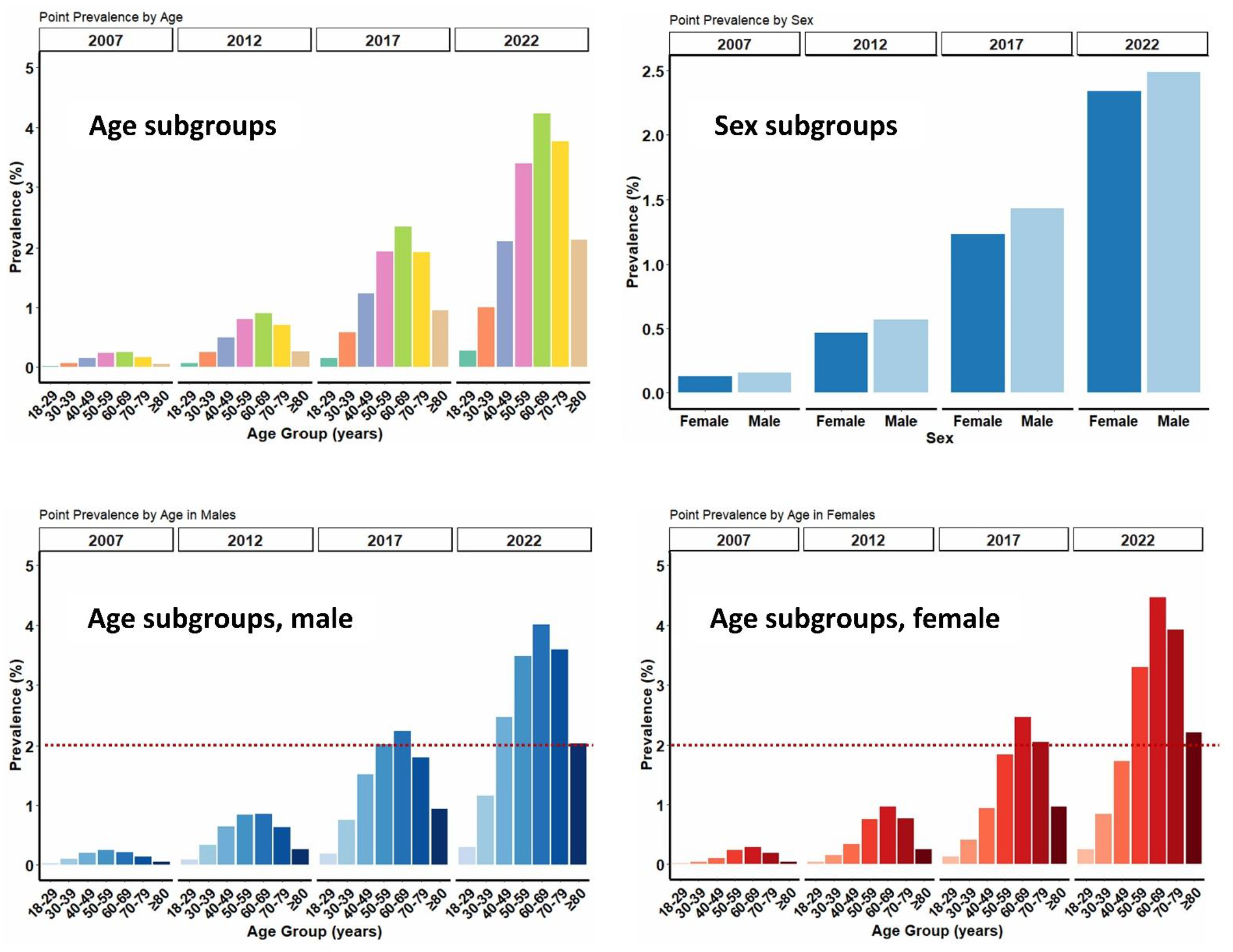
Age and sex subgroups of recorded prevalence of MASLD in the CPRD, in 2007, 2012, 2017, and 2022.

### Increase in incidence of recorded MASLD

The recorded incidence rate of MASLD increased significantly from 2003 (0.09 per 1000 person-years) to 2019, peaking at 3.38 per 1000 person-years (overall trend: p<0.001; **Figure 1**). Following a drop to 2.42 per 1000 person-years in 2020 (likely related to the Sars-CoV-2 pandemic) the incidence rose back to 3.31 in 2022. In the last decade, incidence doubled from 1.60 in 2012 to 3.31 in 2022 per 1000 person-years (p<0.001). Incidence rates increased with age, peaking in the 60–69 age group for males and the 50– 59 age group for females in 2022. Prior to 2022, MASLD incidence was higher in males than females but this reversed with higher incidence in females than in males in 2022 (**Supplementary Figure 2**). A similar trend was observed for annual incidence proportion (**Supplementary Figure 3**).

### Increase in Fib-4 availability in people with MASLD

In the whole cohort, laboratory data were available to calculate the Fib-4 score in 16.8% (N=61,579) of people with MASLD, compared with 9.7% (N=141,882) in matched controls (p<0.001; **Table 1**). The Fib-4 score should be calculated from analytes tested at the same time and over 40% of Fib-4 values were based on results within a one-month (and approximately 80% within a three-month) window around the time of MASLD diagnosis (**Supplementary Figure 4**).

AST was the main limiting factor for calculating Fib-4; it was recorded in only 22.8% (N=83,449) of MASLD cases and 13.7% (N=200,528) of controls, although AST availability in the MASLD cohort increased from 15.2% (N=17,749) pre-2015 to 26.4% (N=65,700) post-2015 (**Supplementary Table 4**). Despite this steady increase, AST availability in incident MASLD cases was <35% in 2022 compared to 85% for ALT. This temporal trend can be observed regardless of ethnicity, with South Asian having lower availability than other ethnicities (**Supplementary Figure 5**). Indeed, the proportion of people with MASLD in whom the Fib-4 score could be calculated (ALT, AST and platelet count all available) increased from 4.3% (N=4,945) pre-2015 to 22.8% (N=56,634) post-2015 (p<0.001). The characteristics of people with MASLD diagnosis were relatively stable before and after 2015 (**Table 2**). After 2015, there were small increases in the proportions of people with MASLD diagnosis who were female, had smoked or had excess alcohol use.

**Table 2.**
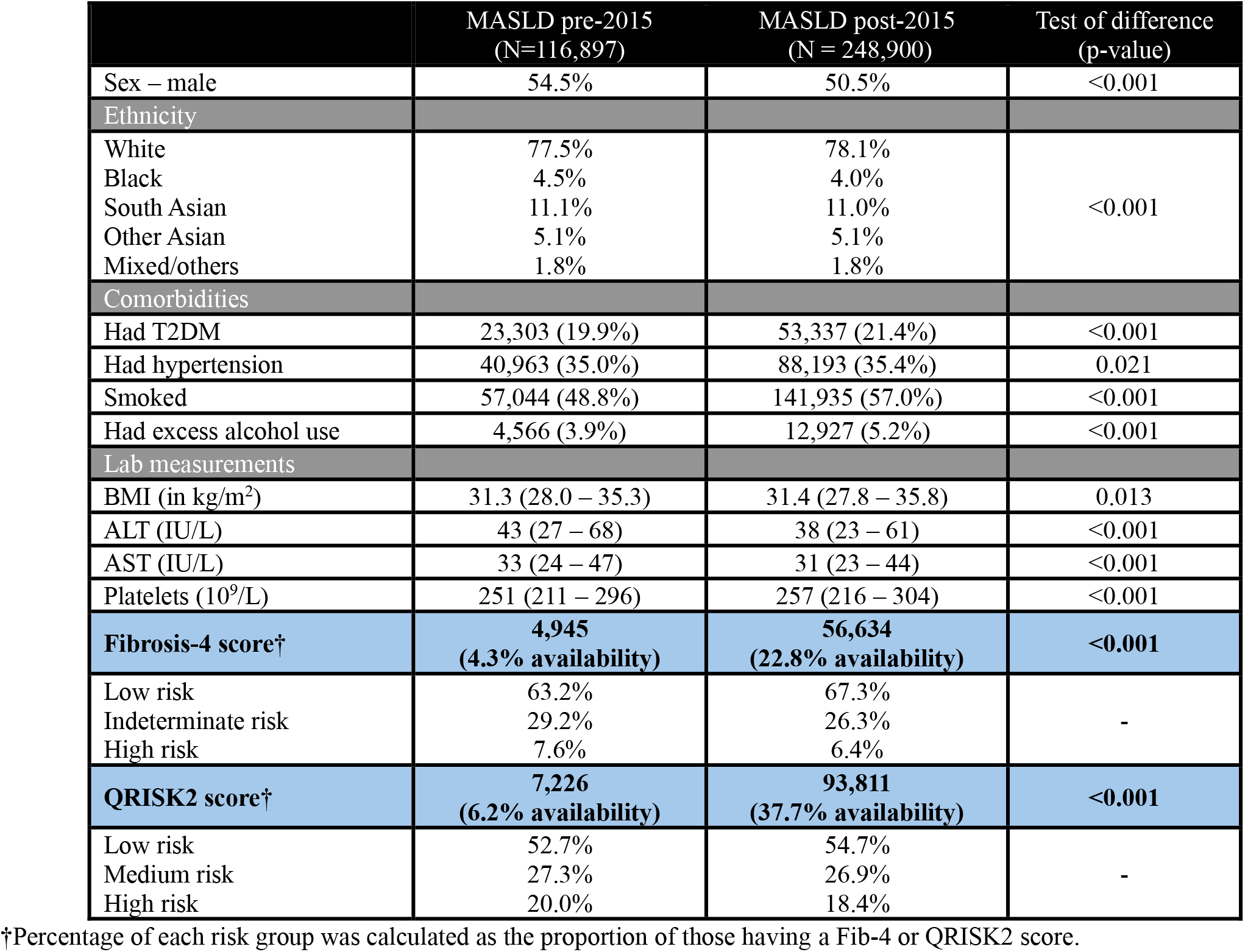
Key descriptive characteristics, Fib-4, and QRISK2 scores of MASLD patients and matched controls, pre-2015 vs post-2015.

In the context of a small fall from pre-2015 to post-2015, ALT (from 43 to 38 IU/L) and AST values (from 33 to 31 IU/L) in MASLD, the proportion of people with MASLD in the high-risk Fib-4 category decreased from 7.6% to 6.4% pre-to post-2015 (**Table 2**). In the same time comparison, availability of the cardiovascular disease risk calculator QRISK2 in people with MASLD diagnosis also rose from 6.2% (N=7,226) pre-2015 to 37.7% (N=93,811) post-2015 (p<0.001; **Table 2**), perhaps indicating greater adoption of risk scoring in clinical care.

### Disparities in fibrosis risk stratification by ethnicity

Ethnicity was more commonly recorded in people with MASLD diagnosis than those without (**Supplementary Table 4**). People of South Asian ethnicity were least likely to have the data available to calculate the Fib-4 score, with an adjusted OR of 0.67 (95% CI: 0.65–0.70; **Table 3A-B**) compared to people of White ethnicity. Among people with a calculable Fib-4, we observed an unexpected distribution of risk. 81.4% (N=3,634) of people of South Asian ethnicity were classified as ‘low risk’, significantly greater than people of White (64.7%; N=27,688) and Black (66.6%; N=1,363) ethnicities. This was despite a significantly higher overall prevalence of T2DM (31.2% [N=9,511] vs 20.8% [N=44,776], p<0.001) in South Asian compared to White ethnicity. While this may relate to a younger median age at MASLD diagnosis than in White patients (46 vs 55 years, p<0.001; **Supplementary Table 5A-B**), and that Fib-4 is said to be less accurate in people aged under 35 years, this is not borne out by sensitivity analysis. After removal of people with available Fib-4 score aged under 35, the proportion of people with low risk scores remained significantly different: 78.2% of South Asian and 61.4% of White ethnicity. In older age, use of age-adjusted cut-off of 2.0 for people aged over 65^16^ increased the proportion of low-risk White patients to 73% (N=31,263) but only marginally increased that in South Asians to 84% (N=3,772) closing this gap (**Table 3C**).

**Table 3.**
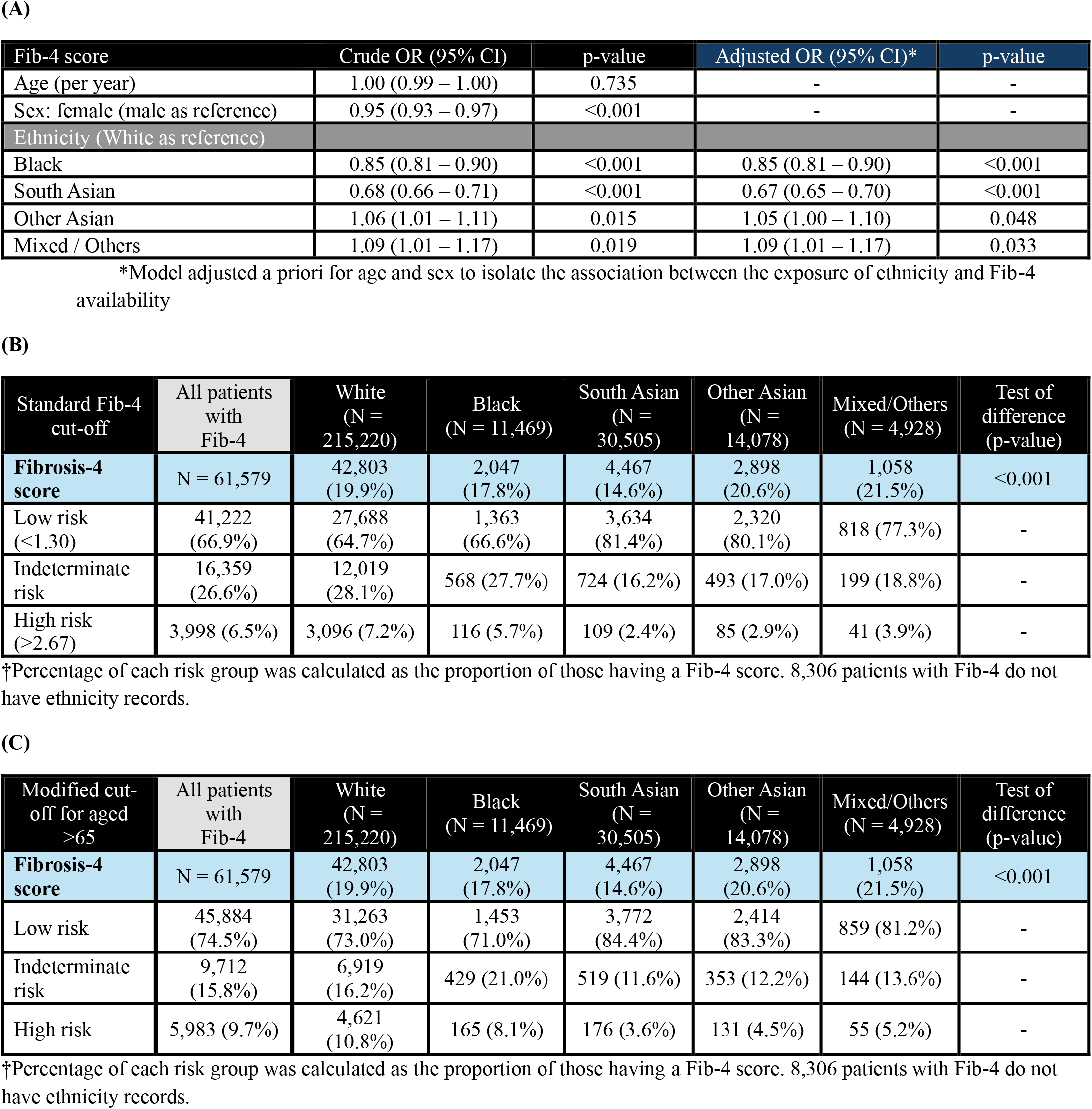
Fibrosis risk stratification statistics. (A) Logistic regression assessing the association between demographic factors and ‘the availability of a calculable Fib-4 score’ in individuals with MASLD (B) Descriptive characteristics of Fib-4 risk counts for each ethnicity. (C) Modified Fib-4 categories using a binary cut-off of low vs high risk for people aged >65.

| Fib-4 score | Crude OR (95% CI) | p-value | Adjusted OR (95% CI)* | p-value |
| --- | --- | --- | --- | --- |
| Age (per year) | 1.00 (0.99 – 1.00) | 0.735 | - | - |
| Sex: female (male as reference) | 0.95 (0.93 – 0.97) | <0.001 | - | - |
| Ethnicity (White as reference) |  |  |  |  |
| Black | 0.85 (0.81 – 0.90) | <0.001 | 0.85 (0.81 – 0.90) | <0.001 |
| South Asian | 0.68 (0.66 – 0.71) | <0.001 | 0.67 (0.65 – 0.70) | <0.001 |
| Other Asian | 1.06 (1.01 – 1.11) | 0.015 | 1.05 (1.00 – 1.10) | 0.048 |
| Mixed / Others | 1.09 (1.01 – 1.17) | 0.019 | 1.09 (1.01 – 1.17) | 0.033 |

\*Model adjusted a priori for age and sex to isolate the association between the exposure of ethnicity and Fib-4 availability

(B)
| Standard Fib-4 cut-off | All patients with Fib-4 | White (N = 215,220) | Black (N = 11,469) | South Asian (N = 30,505) | Other Asian (N = 14,078) | Mixed/Others (N = 4,928) | Test of difference (p-value) |
| --- | --- | --- | --- | --- | --- | --- | --- |
| **Fibrosis-4 score** | N = 61,579 | 42,803 (19.9%) | 2,047 (17.8%) | 4,467 (14.6%) | 2,898 (20.6%) | 1,058 (21.5%) | <0.001 |
| Low risk (<1.30) | 41,222 (66.9%) | 27,688 (64.7%) | 1,363 (66.6%) | 3,634 (81.4%) | 2,320 (80.1%) | 818 (77.3%) | - |
| Indeterminate risk | 16,359 (26.6%) | 12,019 (28.1%) | 568 (27.7%) | 724 (16.2%) | 493 (17.0%) | 199 (18.8%) | - |
| High risk (>2.67) | 3,998 (6.5%) | 3,096 (7.2%) | 116 (5.7%) | 109 (2.4%) | 85 (2.9%) | 41 (3.9%) | - |

†Percentage of each risk group was calculated as the proportion of those having a Fib-4 score. 8,306 patients with Fib-4 do not have ethnicity records.

(C)
| Modified cut-off for aged >65 | All patients with Fib-4 | White (N = 215,220) | Black (N = 11,469) | South Asian (N = 30,505) | Other Asian (N = 14,078) | Mixed/Others (N = 4,928) | Test of difference (p-value) |
| --- | --- | --- | --- | --- | --- | --- | --- |
| **Fibrosis-4 score** | N = 61,579 | 42,803 (19.9%) | 2,047 (17.8%) | 4,467 (14.6%) | 2,898 (20.6%) | 1,058 (21.5%) | <0.001 |
| Low risk | 45,884 (74.5%) | 31,263 (73.0%) | 1,453 (71.0%) | 3,772 (84.4%) | 2,414 (83.3%) | 859 (81.2%) | - |
| Indeterminate risk | 9,712 (15.8%) | 6,919 (16.2%) | 429 (21.0%) | 519 (11.6%) | 353 (12.2%) | 144 (13.6%) | - |
| High risk | 5,983 (9.7%) | 4,621 (10.8%) | 165 (8.1%) | 176 (3.6%) | 131 (4.5%) | 55 (5.2%) | - |

†Percentage of each risk group was calculated as the proportion of those having a Fib-4 score. 8,306 patients with Fib-4 do not have ethnicity records.

## Discussion

This large population study with over 11.7 million individuals’ RWD in the UK shows the evolving epidemiology of recorded MASLD in primary care. Prevalence and incidence have both risen markedly in the last decade, yet despite growing recognition, a diagnostic gap persists. This time-period has also seen a rise in the availability of parameters needed to calculate the most commonly-recommended fibrosis risk score, Fib-4, but availability remains low at approximately one sixth of people with MASLD.

The current study leverages the many benefits of RWD specifically with respect to MASLD including a large-scale, unselected population with comprehensive health records that can be tracked over time^7^. Notwithstanding the challenges of accurately verifying MASLD diagnosis, there remains a major gap between prevalence of MASLD diagnosis (or its proxies) in RWD compared to the estimated prevalence of 25% (95% CI: 21%–31%) in Western Europe from large meta-analyses^1^. A four-nation European RWD study reported a pooled prevalence of 1.85% (95% CI: 0.91%–2.79%)^10^ with similar findings from Copenhagen (0.7% in >100,000 individuals)^17^ and Scotland (1.1% in >130,000 with T2DM)^18^. The current data demonstrate that while UK prevalence has risen substantially to 2.42%, the diagnostic gap remains large compared to expected rates. In order to close this gap, and in keeping with a recent public health target to double the number of individuals diagnosed with MASH^19^, at-risk groups such as those living with T2DM and obesity should be identified and case-finding actively pursued.

The persistent rise in incidence, peaking in 2019, is likely multifactorial and not least reflects the rising prevalence of obesity and T2DM, which are key risk factors of MASLD^20^. However, the scale of the increase, particularly in recent years, also suggests an increase in clinical awareness as obesity and T2DM levels have not risen that fast^21^. The dip in incidence during 2020–2021, almost certainly reflects the disruption of routine primary care services during the COVID-19 pandemic, consistent with lower T2DM incidence and HbA1c monitoring in the NHS during that period^22^.

There was a five-fold increase in Fib-4 score availability before and after 2015 which may reflect increased awareness of liver health. The slight increase after 2015 in people with MASLD diagnoses who have low risk Fib-4 scores may be a further indicator that testing is happening more widely and at on average earlier stages in disease. These findings may reflect impact of initiatives such as the *2016 National Institute for Health and Care Excellence (NICE)* guideline and guidelines from the *European Association for the Study of Liver (EASL)* and the *American Association for the Study of Liver Disease (AASLD)*, which recommend using Fib-4 as first-line testing to risk-stratify MASLD^23,24^. However, increased availability may also reflect the overall increase in testing for metabolic risk factors evidenced in this study by available HbA1c results and the substantial increase in QRISK2 results in people with MASLD. It is important to note that with shared laboratory result reporting, although a result is extracted through the primary care system (as in this study), a test may have originated in secondary care where clinicians may be more inclined to request ‘panels’ of tests with multiple analytes.

The overall proportion of people with analytes available for Fib-4 in this study is lower than other populations with MASLD (42% in Italy and 54% in Spain^10^) or in the general population undergoing routine testing: 22% in France^25^ or 32% Italy^26^. AST is the least available analyte for Fib-4^27^, so AST requesting could be an actionable bottleneck that can be addressed to close this implementation gap. We support efforts to raise awareness of Fib-4, to simplify its requesting and to link the results to accessible, patient-centred pathways of care. It remains to be determined whether and how automatic or ‘reflex’ Fib-4 calculation will improve this pathway^28^. Preliminary 5-year results of the automatic intelligent liver function test (iLFT) have demonstrated broad acceptance in primary care in Tayside, UK^29^. The feasibility and acceptability of such an approach in UK primary care, specifically integrated into the diabetes annual review to overcome these barriers, is currently being evaluated in the prospective PRELUDE1 study^30^.

A major advantage of UK health data is the widespread and relatively detailed recording of ethnicity, which is not available (or permitted) in other territories. Proportionally more people of South Asian ethnicity were in the MASLD group (11.0%) compared to the general UK population (6.9% in 2021^15^) consistent with other reports^31^. South Asian patients were younger and had lower BMI, consistent with increased risk of fibrotic liver disease in this population^32^ and higher levels of BMI-adjusted visceral fat content. Paradoxically, however, this high-risk group were least likely to have the data available to calculate the Fib-4 score. More than 80% of people of South Asian ethnicity were classified as low-risk Fib-4 compared to 65% of people of White ethnicity despite the fact that the former had a much higher proportion of people with T2DM, an important factor for progression to fibrosis. Although the performance of Fib-4 has not been robustly validated in individuals aged under 35 years^16^, the younger age of South Asian patients does not explain this discrepancy. Further studies are needed to examine the possibility that the current scores and thresholds may disadvantage and falsely reassure people of South Asian ethnicity. Indeed, close attention to monitoring of accuracy of Fib-4 as a first-line test in people with T2DM and obesity is also warranted^33^.

The key limitation of this study is that although we followed an internationally-approved coding strategy for MASLD research using EHRs^9^, we cannot ascertain the true proportion of MASH or the stage of fibrosis at diagnosis. It is also not possible to know how and where the MASLD diagnosis was made (imaging, bloods, primary or secondary care) nor the clinical driver for making a diagnosis (symptoms or test results). A consequence of under-diagnosis of MASLD is that it is likely that the 1:4 age- and sex-matched control group contains a large number of people who have the condition but not the diagnostic codes in their health records, thereby diluting the differences described here. We are unable to comment on other, socioeconomic, access or language factors that may have influenced ethnic disparities, which should be the subject for further study. Finally, there is no code in the current dataset for Fib-4, therefore we have used availability of the test results needed to calculate the score as a proxy. Although analytes were largely collected within a short timeframe, it is not possible to know whether the Fib-4 scores were actually calculated by clinicians, or acted upon.

This large real-world population study identifies an increase in MASLD diagnosis and liver fibrosis risk stratification in primary care over the last decade. The diagnostic gap has started to narrow, but more progress is needed to reach public health goals in this disease. Equally urgent is the need for additional research to identify and mitigate any risks of compounding health disparities among different ethnic groups especially in the era of medicines with specific indications in this space.

## Data Availability

All data and analyses were completed and stored in the Data Safe Haven at Queen Mary University of London. No data sharing is available. Access to data can only be granted through researchers approved by Clinical Practice Research Datalink, and within a secure, trusted research environment.

## Contributors

### Author Contributions

*HTH: Conceptualisation, Access to data, Verified the underlying data, Methodology, Analysis, Investigation, Writing – Original Draft, Review & Editing*.

*MH: Methodology, Investigation*.

*WL: Investigation, Writing – Review & Editing.*

*LT: Writing – Review & Editing*.

*NS: Conceptualisation, Writing – Review & Editing*

*WA: Conceptualisation, Access to data, Verified the underlying data, Supervision, Writing – Review & Editing*.

*HTH and WA had access to data and verified the underlying data, and all authors were responsible for the decision to submit for publication*.

## Data sharing statement

All data and analyses were completed and stored in the Data Safe Haven at Queen Mary University of London. No data sharing is available. Access to data can only be granted through researchers approved by CPRD, and within a secure, trusted research environment.

## Acknowledgements

This study is supported in part by a research grant from *Investigator-Initiated Studies Program* of Merck Sharp & Dohme LLC, a subsidiary of Merck & Co., Inc., Rahway, NJ, USA (MISP 101310; WA principal investigator, HTH funded researcher). The interpretation and conclusions contained in this study are those of the author/s alone. The opinions expressed in this paper are those of the authors and do not necessarily represent those of Merck Sharp & Dohme LLC, a subsidiary of Merck & Co., Inc., Rahway, NJ, USA. Barts Charity (MGU0504) sponsored the Multi-Study License of the CPRD at QMUL.

## AI declaration statement

HTH used generative artificial intelligence (Google Gemini 3.1 Pro) as a writing assistant when drafting this manuscript. The manuscript was heavily edited by the authors and verified for accuracy before submission. Authors are accountable for the final content.

