## Supplementary Materials for "Epidemiology, temporal trends, and fibrosis risk stratification in metabolic dysfunction-associated steatotic liver disease in UK primary care: a population-based cohort and nested case-control study"

**Graphical abstract.**………………………………………………………………………………………………………………………………………………………………………………page 2

**Supplementary Table 1. Medical codes of MASLD inclusion and exclusion criteria.**……………………………………………………………………………………….………………page 3

**Supplementary Table 2. Medical codes of comorbidities, ethnicity, and lab measurements.** (A) comorbidities: T2DM, hypertension, any smoking history, and excess alcohol use. (B) Ethnicity. (C) BMI and lab measurements..………………………………………………………………………………………………………………………………………………………………..…page 6

**Supplementary Table 3. Characteristics of MASLD vs MASLD excluding patients with any ‘excess alcohol use’ record.**……………………………………………………………page 24

**Supplementary Table 4. The availability of characteristics and lab measurements of MASLD patients and matched controls in the CPRD, and pre-2015 vs post-2015.**………...page 25

**Supplementary Table 5. Descriptive characteristics of MASLD patients between White Caucasians and South Asians**. (A) All individuals. (B) Individuals with low risk Fib-4 (Fib-4 <1.30). For categorical variables, records were reported as counts (in percentages); for continuous variables, records were reported as medians (with IQRs). ……………………………………...page 26

**Supplementary Figure 1. The study flow chart.**……………………………………………………………………………………………………………………………………………...page 28

**Supplementary Figure 2. Age and sex subgroups of recorded incidence of MASLD in the CPRD, in 2007, 2012, 2017, and 2022.**……………………………………………………page 29

**Supplementary Figure 3. Annual incidence proportion of MASLD between 2003 and 2022, and age and sex subgroups in 2007, 2012, 2017, and 2022.**…………………………..page 30

**Supplementary Figure 4. Distribution of time points for each Fib-4 component in MASLD patients.**…………………………………………………………………………………..page 31

**Supplementary Figure 5. Temporal trend of Fib-4 components availability at the time of MASLD diagnosis, 2003 to 2022.** An effective test is defined as the closest record within [-2 years, +6 months] of the MASLD index date. (A) Overall trend. (B) Racial-ethnic trend. …………………………………………………………………………………………………………….page 32

**Graphical abstract.**
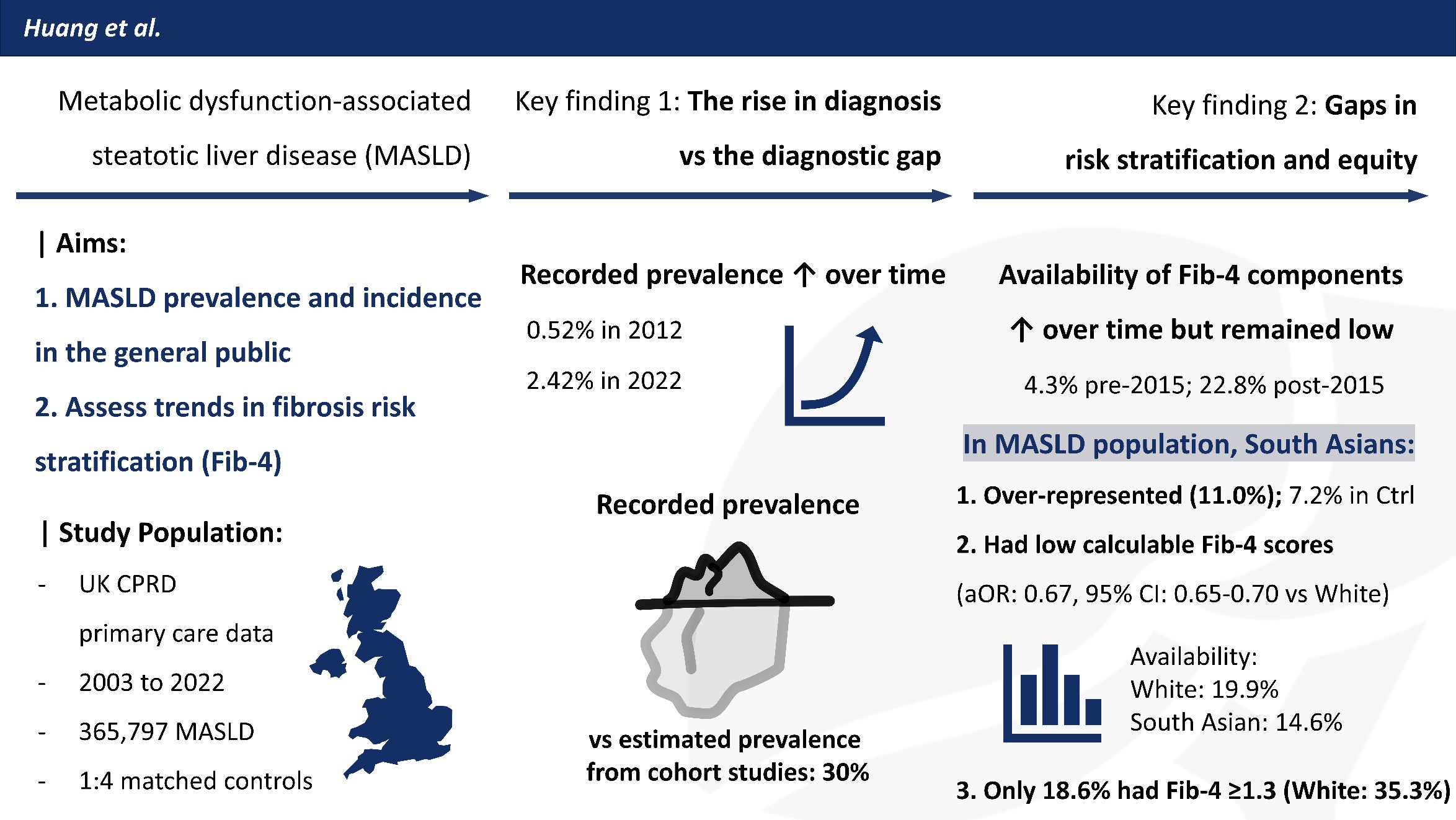

**Supplementary Table 1. Medical codes of MASLD inclusion and exclusion criteria.**

|  | **Read Code** | **Term** | **SNOMED CT Code** |
| --- | --- | --- | --- |
| **MASLD inclusion** | J61y800 | Nonalcoholic steatohepatitis | 442685003 |
|  | J61y.00 | Chronic nonalcoholic liver disease | 79720007 |
|  | J61y911 | Fatty liver | 197321007 |
|  | J61y100 | Non-alcoholic fatty liver | 197315008 |
|  | J61y900 | Fatty change of liver | 197321007 |
|  | J61y700 | Steatosis of liver | 197321007 |
|  | Jyu7200 | [X]Other specified inflammatory liver diseases | 128241005 |
|  | J61yz00 | Other non-alcoholic chronic liver disease NOS | 328383001 |
| **Exclusion criteria** | A70z000 | Hepatitis C | 50711007 |
|  | J616000 | Primary biliary cirrhosis | 31712002 |
|  | J63B.00 | Autoimmune hepatitis | 408335007 |
|  | 43X3.00 | Hepatitis C antibody test positive | 314706002 |
|  | 43B4.00 | Hepatitis B surface antigen positive | 165806002 |
|  | A70..00 | Viral hepatitis | 3738000 |
|  | A703.00 | Serum hepatitis | 66071002 |
|  | A707300 | Chronic viral hepatitis B | 61977001 |
|  | A705000 | Viral hepatitis C | 50711007 |
|  | - | Haemochromatosis | 399187006 |
|  | A707200 | Chronic hepatitis C | 128302006 |
|  | C376200 | alpha-1-Antitrypsin deficiency | 30188007 |
|  | 4J3B.00 | Hepatitis C viral load | 992751000000109 |
|  | J661700 | Primary sclerosing cholangitis | 197441003 |
|  | 141E.00 | History of hepatitis B | 429721005 |
|  | 9kZ..11 | Hepatitis B screening positive | 356121000000100 |
|  | J614111 | Autoimmune chronic active hepatitis | 197284004 |
|  | 43jG.00 | Hepatitis B nucleic acid detection | 1031571000000101 |
|  | 4JQ3.00 | Hepatitis C virus genotype | 992771000000100 |
|  | 43j5.00 | Hepatitis C nucleic acid detection | 1030511000000102 |
|  | 4J3D.00 | Hepatitis B viral load | 222531000000101 |
|  | C351011 | Wilson's disease | 88518009 |
|  | 9kV..11 | Hepatitis C screening positive | 362751000000101 |
|  | 4JQD.11 | Hepatitis C PCR positive | 760421000000100 |
|  | A70C.00 | Hepatitis C genotype 3 | 824871000000104 |
|  | A70A.00 | Hepatitis C genotype 1 | 824841000000105 |
|  | 9NgR.00 | On hepatitis C treatment plan | 829731000000106 |
|  | A707.00 | Chronic viral hepatitis | 10295004 |
|  | A707100 | Chronic viral hepatitis B without delta-agent | 186639003 |
|  | 4JQF.00 | Hepatitis C antigen positive | 812181000000106 |
|  | 9kZ..00 | Hepatitis B screening positive - enhanced services administration | 356121000000100 |
|  | 4JQD.00 | Hepatitis C viral ribonucleic acid polymerase chain reaction positive | 760421000000100 |
|  | 43B5.00 | Hepatitis e antigen present | 165807006 |
|  | 9kV..00 | Hepatitis C screening positive - enhanced services admin | 362751000000101 |
|  | Q409100 | Congenital viral hepatitis B infection | 60498001 |
|  | 14i..00 | H/O hepatitis C antiviral drug therapy | 829691000000100 |
|  | A707000 | Chronic viral hepatitis B with delta-agent | 235869004 |
|  | - | Primary biliary cholangitis | 31712002 |
|  | A703.99 | Serum hepatitis | 66071002 |
|  | 8BB5.00 | 12 week virologic response to hepatitis C treatment | 829241000000108 |
|  | C376100 | Alpha-1-antitrypsin hepatitis | 190944000 |
|  | A705100 | Acute delta-(super)infection of hepatitis B carrier | 235865005 |
|  | 9kR..00 | Chronic hepatitis annual review - enhanced services admin | 362421000000102 |
|  | A70B.00 | Hepatitis C genotype 2 | 824851000000108 |
|  | A70D.00 | Hepatitis C genotype 4 | 824881000000102 |
|  | A707X00 | Chronic viral hepatitis, unspecified | 10295004 |
|  | AyuB200 | [X]Chronic viral hepatitis, unspecified | 10295004 |
|  | AyuB100 | [X]Other chronic viral hepatitis | 10295004 |
|  | 9kX..00 | Hepatitis status 6 months post treatment - enhanced serv adm | 355741000000101 |
|  | A70F.00 | Hepatitis C genotype 6 | 824901000000104 |
|  | A70E.00 | Hepatitis C genotype 5 | 824891000000100 |

- **Reference:**
- Alexander M, Loomis AK, Fairburn-Beech J *et al.* Real-world data reveal a diagnostic gap in non-alcoholic fatty liver disease. *BMC Med* **16**, 130 (2018). <https://doi.org/10.1186/s12916-018-1103-x>
- Kuan V, Denaxas S, Gonzalez-Izquierdo A *et al.* *PH111 / 222 - Autoimmune liver disease*. Phenotype Library [Online]. 06 October 2021. Available from: <http://phenotypes.healthdatagateway.org/phenotypes/PH111/version/222/detail/>
- Kuan V, Denaxas S, Gonzalez-Izquierdo A *et al.* *PH137 / 274 - Chronic viral hepatitis*. Phenotype Library [Online]. 06 October 2021. Available from: <http://phenotypes.healthdatagateway.org/phenotypes/PH137/version/274/detail/>

**Supplementary Table 2. Medical codes of comorbidities, ethnicity, and lab measurements.** (A) comorbidities: T2DM, hypertension, any smoking history, and excess alcohol use. (B) Ethnicity. (C) BMI and lab measurements.

|  | **Read Code** | **Term** | **SNOMED CT Code** |
| --- | --- | --- | --- |
| **T2DM** | C10F.00 | Type 2 diabetes mellitus | 44054006 |
|  | 66At100 | Type II diabetic dietary review | 754121000000107 |
|  | C109.00 | Non-insulin dependent diabetes mellitus | 44054006 |
|  | C10FJ00 | Insulin treated Type 2 diabetes mellitus | 237599002 |
|  | C10F.11 | Type II diabetes mellitus | 44054006 |
|  | 66Ao.00 | Diabetes type 2 review | 279321000000104 |
|  | C109.12 | Type 2 diabetes mellitus | 44054006 |
|  | C100112 | Non-insulin dependent diabetes mellitus | 44054006 |
|  | C10FM00 | Persistent microalbuminuria associated with type 2 diabetes mellitus | 420715001 |
|  | C109J00 | Insulin treated Type 2 diabetes mellitus | 237599002 |
|  | C109.13 | Type II diabetes mellitus | 44054006 |
|  | C109.11 | NIDDM - Non-insulin dependent diabetes mellitus | 44054006 |
|  | C10FL00 | Persistent proteinuria associated with type 2 diabetes mellitus | 421986006 |
|  | C109700 | Type II diabetes mellitus uncontrolled | 443694000 |
|  | C10F700 | Type II diabetes mellitus poorly controlled | 443694000 |
|  | 66At111 | Type 2 diabetic dietary review | 754121000000107 |
|  | C10F900 | Type 2 diabetes mellitus without complication | 313436004 |
|  | C10FC00 | Diabetes type 2 with nephropathy | 420279001 |
|  | C10F600 | Retinopathy with type 2 diabetes mellitus | 422034002 |
|  | C109900 | Non-insulin-dependent diabetes mellitus without complication | 313436004 |
|  | C10FR00 | Gastroparesis with type 2 diabetes mellitus | 713703005 |
|  | C10F000 | Renal disorder associated with type II diabetes mellitus | 420279001 |
|  | C10F300 | Type 2 diabetes mellitus with multiple complications | 190388001 |
|  | C10FN00 | Ketoacidosis in type 2 diabetes mellitus | 421750000 |
|  | C10FM11 | Type II diabetes mellitus with persistent microalbuminuria | 420715001 |
|  | C10F200 | Neurologic disorder associated with type 2 diabetes mellitus | 421326000 |
|  | C10FJ11 | Insulin treated Type II diabetes mellitus | 237599002 |
|  | C109J12 | Insulin treated Type II diabetes mellitus | 237599002 |
|  | C10F100 | Disorder of eye with type 2 diabetes mellitus | 422099009 |
|  | C10FQ00 | Exudative maculopathy associated with type 2 diabetes mellitus | 421779007 |
|  | C109712 | Type 2 diabetes mellitus - poor control | 443694000 |
|  | C10FK00 | Hyperosmolar non-ketotic state in type 2 diabetes mellitus | 395204000 |
|  | C109J11 | Insulin treated non-insulin dependent diabetes mellitus | 237599002 |
|  | C10F400 | Type 2 diabetes mellitus with ulcer | 190389009 |
|  | C10F911 | Type II diabetes mellitus without complication | 313436004 |
|  | C109C11 | Renal disorder due to type 2 diabetes mellitus | 420279001 |
|  | C10FB00 | Polyneuropathy due to type 2 diabetes mellitus | 713706002 |
|  | C10FH00 | Type 2 diabetes mellitus with neuropathic arthropathy | 314904008 |
|  | C109711 | Type II diabetes mellitus - poor control | 443694000 |
|  | C10D.00 | Diabetes mellitus autosomal dominant type II | 237604008 |
|  | C109C12 | Type 2 diabetes mellitus with nephropathy | 420279001 |
|  | C109000 | Non-insulin-dependent diabetes mellitus with renal comps | 420279001 |
|  | C109600 | Non-insulin-dependent diabetes mellitus with retinopathy | 422034002 |
|  | C109100 | Non-insulin-dependent diabetes mellitus with ophthalm comps | 422099009 |
|  | C10FL11 | Type II diabetes mellitus with persistent proteinuria | 421986006 |
|  | C109200 | Non-insulin-dependent diabetes mellitus with neuro comps | 421326000 |
|  | C10FE00 | Type 2 diabetes mellitus with diabetic cataract | 420756003 |
|  | C109K00 | Hyperosmolar non-ketotic state in type 2 diabetes mellitus | 395204000 |
|  | C10FR11 | Type II diabetes mellitus with gastroparesis | 713703005 |
|  | C10FF00 | Type 2 diabetes mellitus with peripheral angiopathy | 314902007 |
|  | C109400 | Non-insulin-dependent diabetes mellitus with ulcer | 190389009 |
|  | C10FA00 | Type 2 diabetes mellitus with mononeuropathy | 420436000 |
|  | C10F711 | Type II diabetes mellitus - poor control | 443694000 |
|  | C109300 | Non-insulin-dependent diabetes mellitus with multiple complications | 190388001 |
|  | C10F500 | Type 2 diabetes mellitus with gangrene | 421631007 |
|  | C10FQ11 | Exudative maculopathy with type 2 diabetes mellitus | 421779007 |
|  | C10FD00 | Type 2 diabetes mellitus with hypoglycaemic coma | 719216001 |
|  | C10FG00 | Type 2 diabetes mellitus with arthropathy | 314903002 |
|  | C109012 | Type 2 diabetes mellitus with renal complications | 420279001 |
|  | C109C00 | Non-insulin dependent diabetes mellitus with nephropathy | 420279001 |
|  | C109112 | Type 2 diabetes mellitus with ophthalmic complications | 422099009 |
|  | C109312 | Type 2 diabetes mellitus with multiple complications | 190388001 |
|  | C109611 | Type II diabetes mellitus with retinopathy | 422034002 |
|  | C109E11 | Cataract due to diabetes mellitus type 2 | 420756003 |
|  | C109612 | Type 2 diabetes mellitus with retinopathy | 422034002 |
|  | C109E00 | Non-insulin depend diabetes mellitus with diabetic cataract | 420756003 |
|  | C109212 | Type 2 diabetes mellitus with neurological complications | 421326000 |
|  | C109500 | Gangrene associated with type 2 diabetes mellitus | 421631007 |
|  | C10F611 | Type II diabetes mellitus with retinopathy | 422034002 |
|  | C109E12 | Type 2 diabetes mellitus with diabetic cataract | 420756003 |
|  | C10FE11 | Type II diabetes mellitus with diabetic cataract | 420756003 |
|  | C109412 | Type 2 diabetes mellitus with ulcer | 190389009 |
|  | C10FC11 | Type II diabetes mellitus with nephropathy | 420279001 |
|  | C109912 | Type 2 diabetes mellitus without complication | 313436004 |
|  | C10FK11 | Hyperosmolar non-ketotic state in type II diabetes mellitus | 395204000 |
|  | C109F00 | Non-insulin-dependent diabetes mellitus with peripheral angiopathy | 314902007 |
|  | C10FN11 | Type II diabetes mellitus with ketoacidosis | 421750000 |
|  | C109911 | Type II diabetes mellitus without complication | 313436004 |
|  | C109011 | Type II diabetes mellitus with renal complications | 420279001 |
|  | C10FD11 | Hypoglycaemic coma co-occurrent and due to diabetes mellitus type II | 719216001 |
|  | C10FF11 | Type II diabetes mellitus with peripheral angiopathy | 314902007 |
|  | C109411 | Type II diabetes mellitus with ulcer | 190389009 |
|  | C109D00 | Non-insulin dependent diabetes mellitus with hypoglyca coma | 719216001 |
|  | C109B00 | Non-insulin dependent diabetes mellitus with polyneuropathy | 713706002 |
|  | C10FG11 | Type II diabetes mellitus with arthropathy | 314903002 |
|  | C109211 | Neurological disorder with diabetes type 2 | 421326000 |
|  | C109111 | Disorder of eye with type 2 diabetes mellitus | 422099009 |
|  | C10FP00 | Ketoacidotic coma in type 2 diabetes mellitus | 421847006 |
|  | C10FH11 | Type II diabetes mellitus with neuropathic arthropathy | 314904008 |
|  | C10FA11 | Type II diabetes mellitus with mononeuropathy | 420436000 |
|  | C10FB11 | Type II diabetes mellitus with polyneuropathy | 713706002 |
|  | C10F311 | Type II diabetes mellitus with multiple complications | 190388001 |
|  | C109A00 | Non-insulin dependent diabetes mellitus with mononeuropathy | 420436000 |
|  | C10F411 | Type II diabetes mellitus with ulcer | 190389009 |
|  | C109A11 | Mononeuropathy with type 2 diabetes mellitus | 420436000 |
|  | C10F011 | Type II diabetes mellitus with renal complications | 420279001 |
|  | C109D12 | Type 2 diabetes mellitus with hypoglycaemic coma | 719216001 |
|  | C109B11 | Type II diabetes mellitus with polyneuropathy | 713706002 |
|  | C109H12 | Type 2 diabetes mellitus with neuropathic arthropathy | 314904008 |
|  | C109H00 | Non-insulin dependent diabetes mellitus with neuropathic arthropathy | 314904008 |
|  | C109F11 | Type II diabetes mellitus with peripheral angiopathy | 314902007 |
|  | C109A12 | Type 2 diabetes mellitus with mononeuropathy | 420436000 |
|  | C109512 | Type 2 diabetes mellitus with gangrene | 421631007 |
|  | C10F211 | Type II diabetes mellitus with neurological complications | 421326000 |
|  | C109H11 | Type II diabetes mellitus with neuropathic arthropathy | 314904008 |
|  | C109G12 | Type 2 diabetes mellitus with arthropathy | 314903002 |
|  | C109B12 | Type 2 diabetes mellitus with polyneuropathy | 713706002 |
|  | C10F111 | Type II diabetes mellitus with ophthalmic complications | 422099009 |
|  | C10F511 | Type II diabetes mellitus with gangrene | 421631007 |
|  | C109G00 | Non-insulin dependent diabetes mellitus with arthropathy | 314903002 |
|  | C109F12 | Type 2 diabetes mellitus with peripheral angiopathy | 314902007 |
|  | C109511 | Type II diabetes mellitus with gangrene | 421631007 |
|  | C109311 | Type II diabetes mellitus with multiple complications | 190388001 |
|  | C10FP11 | Ketoacidotic coma in type II diabetes mellitus | 421847006 |
|  | C109D11 | Type II diabetes mellitus with hypoglycaemic coma | 719216001 |
|  | C109G11 | Type II diabetes mellitus with arthropathy | 314903002 |
| **Hypertension** | G20..00 | Essential hypertension | 59621000 |
|  | G2...00 | Hypertensive disease | 38341003 |
|  | 662d.00 | Hypertension annual review | 401118009 |
|  | G20z.11 | Hypertension | 38341003 |
|  | 662c.00 | Hypertension six month review | 401048005 |
|  | G20z.00 | Essential hypertension NOS | 59621000 |
|  | G2z..00 | Hypertensive disorder | 38341003 |
|  | 8BL0.00 | Patient on maximal tolerated antihypertensive therapy | 407567007 |
|  | 14A2.00 | H/O: hypertension | 161501007 |
|  | G201.00 | Benign essential hypertension | 1201005 |
|  | 662O.00 | On treatment for hypertension | 302192008 |
|  | 662G.00 | Hypertensive treatment changed | 170587004 |
|  | 8B26.00 | Antihypertensive therapy | 308116003 |
|  | G2...11 | BP - hypertensive disease | 38341003 |
|  | G202.00 | Systolic hypertension | 56218007 |
|  | 662F.00 | Treatment for hypertension started | 170586008 |
|  | G28..00 | Stage 2 hypertension (NICE - National Institute for Health and Clinical Excellence 2011) | 846371000000103 |
|  | G25..00 | Stage 1 hypertension (NICE - National Institute for Health and Clinical Excellence 2011) | 843821000000102 |
|  | 212K.00 | Hypertension resolved | 162659009 |
|  | G24..00 | Secondary hypertension | 31992008 |
|  | G21..00 | Hypertensive heart disease | 64715009 |
|  | G250.00 | Stage 1 hypertension (NICE 2011) without evidence of end organ damage | 908631000000108 |
|  | G22..00 | Hypertensive renal disease | 38481006 |
|  | G200.00 | Malignant essential hypertension | 78975002 |
|  | G25..11 | Stage 1 hypertension | 843821000000102 |
|  | F421300 | Hypertensive retinopathy | 6962006 |
|  | 662b.00 | Moderate hypertension control | 401117004 |
|  | G22z.11 | Renal hypertension | 38481006 |
|  | G2y..00 | Other specified hypertensive disease | 38341003 |
|  | G203.00 | Diastolic hypertension | 48146000 |
|  | 8I3N.00 | Hypertension treatment refused | 401047000 |
|  | G21z011 | Cardiomegaly - hypertensive | 275516004 |
|  | G251.00 | Stage 1 hypertension (NICE 2011) with evidence of end organ damage | 908651000000101 |
|  | G672.00 | Hypertensive encephalopathy | 50490005 |
|  | G26..11 | Severe hypertension | 843841000000109 |
|  | G22z.00 | Hypertensive renal disease NOS | 38481006 |
|  | G222.00 | Hypertensive renal disease with renal failure | 49220004 |
|  | G24z000 | Renovascular hypertension | 123799005 |
|  | L127000 | Pre-eclampsia or eclampsia with hypertension unspecified | 198997005 |
|  | G20..12 | Primary hypertension | 59621000 |
|  | TJC7.00 | Adverse reaction to other antihypertensives | 293495006 |
|  | G244.00 | Hypertension secondary to endocrine disorders | 194788005 |
|  | TJC7z00 | Antihypertensive adverse reaction | 293495006 |
|  | G24z.00 | Secondary hypertension NOS | 31992008 |
|  | G24zz00 | Secondary hypertension NOS | 31992008 |
|  | G672.11 | Hypertensive crisis | 50490005 |
|  | G220.00 | Malignant hypertensive renal disease | 65443008 |
|  | G200.99 | Malignant hypertension | 78975002 |
|  | G241000 | Secondary benign renovascular hypertension | 73410007 |
|  | G240.00 | Malignant secondary hypertension | 89242004 |
|  | G21z100 | Hypertensive heart disease NOS with CCF | 64715009 |
|  | G221.00 | Benign hypertensive renal disease | 193003 |
|  | G23..00 | Hypertensive heart AND renal disease | 86234004 |
|  | G21z.00 | Hypertensive heart disease NOS | 64715009 |
|  | G24z100 | Hypertension secondary to drug | 194791005 |
|  | G241.00 | Secondary benign hypertension | 194785008 |
|  | G211.00 | Benign hypertensive heart disease | 36221001 |
|  | G211100 | Benign hypertensive heart disease with congestive cardiac failure | 194767001 |
|  | G210.00 | Malignant hypertensive heart disease | 54225002 |
|  | G21zz00 | Hypertensive heart disease NOS | 64715009 |
|  | L127z00 | Pre-eclampsia or eclampsia + pre-existing hypertension NOS | 198997005 |
|  | G21z000 | Hypertensive heart disease NOS without CCF | 64715009 |
|  | Gyu2.00 | [X]Hypertensive diseases | 38341003 |
|  | Gyu2100 | [X]Hypertension secondary to other renal disorders | 31992008 |
|  | G241z00 | Benign secondary hypertension | 194785008 |
|  | U60C51A | [X] Adverse reaction to antihypertensives NOS | 293495006 |
|  | F404200 | Blind hypertensive eye | 264008 |
|  | G240z00 | Secondary malignant hypertension NOS | 89242004 |
|  | U60C500 | [X]Oth antihyperten drug caus advers eff in therap use, NEC | 293495006 |
|  | G233.00 | Hypertensive heart and renal disease with renal failure | 194780003 |
|  | G211000 | Benign hypertensive heart disease without CCF | 77970009 |
|  | Gyu2000 | [X]Other secondary hypertension | 31992008 |
|  | G232.00 | Hypertensive heart and renal disease with (congestive) heart failure | 194779001 |
|  | L122000 | Other pre-existing hypertension in preg/childb/puerp unspec | 86041002 |
|  | G230.00 | Malignant hypertensive heart AND renal disease | 66610008 |
|  | L128.00 | Pre-existing hypertension complicating pregnancy, childbirth and puerperium | 199005000 |
|  | G210z00 | Malignant hypertensive heart disease NOS | 54225002 |
|  | G240000 | Secondary malignant renovascular hypertension | 194783001 |
|  | G210000 | Malignant hypertensive heart disease without congestive heart failure | 36315003 |
|  | U60C511 | [X] Adverse reaction to other antihypertensives | 293495006 |
|  | L128200 | Pre-existing secondary hypertension complicating pregnancy, childbirth and puerperium | 199008003 |
|  | L122100 | Other pre-existing hypertension in preg/childb/puerp - deliv | 86041002 |
|  | 662r.00 | Trial withdrawal of antihypertensive therapy | 299561000000105 |
|  | L122z00 | Other pre-existing hypertension in preg/childb/puerp NOS | 86041002 |
|  | G210100 | Malignant hypertensive heart disease with congestive cardiac failure | 83105008 |
|  | G234.00 | Hypertensive heart and renal disease with both (congestive) heart failure and renal failure | 194781004 |
|  | G23..99 | Hypertensive renal + heart dis | 86234004 |
|  | L122300 | Other pre-exist hypertension in preg/childb/puerp-not deliv | 86041002 |
|  | G23z.00 | Hypertensive heart and renal disease NOS | 86234004 |
|  | G231.00 | Benign hypertensive heart AND renal disease | 66052004 |
|  | G211z00 | Benign hypertensive heart disease NOS | 36221001 |
|  | 7Q01.00 | High cost hypertension drugs | 220901000000101 |
|  | L128000 | Pre-existing hypertensive heart disease complicating pregnancy, childbirth and the puerperium | 199006004 |
|  | L128100 | Pre-existing hypertensive heart and renal disease complicating pregnancy, childbirth and the puerperium | 199007008 |
| **Any smoking history** | 137S.00 | Ex-smoker | 8517006 |
|  | 137P.00 | Cigarette smoker | 65568007 |
|  | 137R.00 | Current smoker | 77176002 |
|  | 1374.00 | Moderate cigarette smoker (10-19 cigs/day) | 160604004 |
|  | 1373.00 | Light cigarette smoker (1-9 cigs/day) | 160603005 |
|  | 1379.00 | Ex-moderate cigarette smoker (10-19/day) | 266923002 |
|  | 137G.00 | Trying to give up smoking | 160616005 |
|  | 137..00 | Tobacco smoking consumption | 266918002 |
|  | 137j.00 | Ex-cigarette smoker | 281018007 |
|  | 137P.11 | Smoker | 77176002 |
|  | 1378.00 | Ex-light cigarette smoker (1-9/day) | 266922007 |
|  | 1375.00 | Heavy cigarette smoker (20-39 cigs/day) | 160605003 |
|  | 137F.00 | Ex-smoker - amount unknown | 8517006 |
|  | 137A.00 | Ex-heavy cigarette smoker (20-39/day) | 266924008 |
|  | 1377.00 | Ex-trivial cigarette smoker (<1/day) | 266921000 |
|  | 1372.00 | Trivial cigarette smoker (less than one cigarette/day) | 266920004 |
|  | 137d.00 | Not interested in stopping smoking | 394873005 |
|  | - | Tobacco smoking behaviour - finding | 365981007 |
|  | 137J.00 | Cigar smoker | 59978006 |
|  | 137H.00 | Pipe smoker | 82302008 |
|  | 137B.00 | Ex-very heavy cigarette smoker (40+/day) | 266925009 |
|  | 1372.11 | Occasional smoker | 428041000124106 |
|  | 1376.00 | Very heavy cigarette smoker (40+ cigs/day) | 160606002 |
|  | 137..11 | Smoker - amount smoked | 266918002 |
|  | 137m.00 | Failed attempt to stop smoking | 446172000 |
|  | 137O.00 | Ex-cigar smoker | 160621008 |
|  | 137N.00 | Ex-pipe smoker | 160620009 |
|  | - | Ex-Cigarette Smoker | 649861000006105 |
|  | 137l.00 | Ex roll-up cigarette smoker | 492191000000103 |
|  | 137K000 | Recently stopped smoking | 517211000000106 |
|  | 137e.00 | Smoking restarted | 308438006 |
|  | - | Ex- Rolled Tobacco Smoker | 649851000006108 |
|  | - | Current Smoker NOS | 604961000006105 |
|  | - | Cigarette smoker | 854021000006104 |
|  | - | Smoker (Read codes) | 137711000006107 |
|  | - | Smokes tobacco daily | 449868002 |
|  | 137F.99 | EX-Smoker NOS | 266928006 |
|  | - | Light cigarette smoker | 230060001 |
|  | - | Occasional cigarette smoker | 230059006 |
|  | - | Ex-cigarette smoker amount unknown | 266928006 |
|  | - | Ex-smoker for more than 1 year | 48031000119106 |
|  | - | Moderate cigarette smoker | 230062009 |
|  | - | Heavy cigarette smoker | 230063004 |
|  | - | Grade C moderate smoker (11-20/day) | 854981000006101 |
|  | - | Smokes/uses tobacco products | 961581000006105 |
|  | - | Tobacco smoking consumption unknown | 266927001 |
|  | - | Grade B light smoker (1-10/day) | 854961000006106 |
|  | - | Current smoker | 854071000006103 |
|  | - | Primary carer current smoker | 1888831000006106 |
|  | - | Exposure to cigarette/cigar smoke | 981841000006104 |
|  | - | Occasional tobacco smoker | 428041000124106 |
|  | - | Grade D heavy smoker (>20 Day) | 855001000006105 |
|  | - | Gradual smoking reduction | 852111000006102 |
|  | - | Ex-light smoker (1-9/day) | 1092111000000104 |
|  | - | Ex-moderate smoker (10-19/day) | 1092091000000109 |
|  | - | Ex-heavy smoker (20-39/day) | 1092071000000105 |
|  | - | Ex-trivial smoker (<1/day) | 1092131000000107 |
|  | - | Moderate smoker (20 or less per day) | 56578002 |
|  | - | Heavy smoker (over 20 per day) | 56771006 |
|  | - | Ex-smoker for less than 1 year | 735128000 |
| **Excess Alcohol Use** | E23..00 | Alcohol dependence syndrome | 66590003 |
|  | E23..11 | Alcoholism | 66590003 |
|  | E250.00 | Nondependent alcohol abuse | 268645007 |
|  | 136S.00 | Hazardous alcohol use | 198421000000108 |
|  | E231000 | Alcohol dependence | 66590003 |
|  | E23z.00 | Alcohol dependence syndrome NOS | 66590003 |
|  | J612.00 | Alcoholic cirrhosis of liver | 420054005 |
|  | E01y000 | Alcohol withdrawal syndrome | 191480000 |
|  | J613.00 | Alcoholic liver damage | 41309000 |
|  | 8H7p.00 | Referral to community alcohol team | 390857005 |
|  | 136T.00 | Harmful alcohol use | 198431000000105 |
|  | 8HHe.00 | Referral to community drug and alcohol team | 417096006 |
|  | E231.00 | Chronic alcoholism | 66590003 |
|  | 1462.00 | H/O: alcoholism | 161466001 |
|  | 136R.00 | Binge drinker | 228315001 |
|  | 136W.00 | Alcohol misuse | 15167005 |
|  | J610.00 | Alcoholic fatty liver | 50325005 |
|  | E250000 | Nondependent alcohol abuse, unspecified | 268645007 |
|  | J153.00 | Alcoholic gastritis | 2043009 |
|  | 8HkG.00 | Referral to specialist alcohol treatment service | 431260004 |
|  | Eu10100 | Alcohol abuse | 15167005 |
|  | J617.00 | Alcoholic hepatitis | 235875008 |
|  | Eu10800 | Alcohol withdrawal-induced seizure | 308742005 |
|  | ZV6D600 | Alcoholism counselling | 24165007 |
|  | ZV4KC00 | [V] Alcohol use | 219006 |
|  | 8IEA.00 | Referral to community alcohol team declined | 781191000000101 |
|  | 66e..00 | Alcohol disorder monitoring | 413130000 |
|  | Eu10.00 | Alcohol-induced organic mental disorder | 29212009 |
|  | 66e0.00 | Alcohol abuse monitoring | 247721000000109 |
|  | Eu10211 | [X]Alcohol addiction | 66590003 |
|  | J671000 | Alcohol-induced chronic pancreatitis | 235952002 |
|  | G555.00 | Alcoholic cardiomyopathy | 83521008 |
|  | J611.00 | Acute alcoholic hepatitis | 9953008 |
|  | Eu10200 | [X]Mental and behav dis due to use alcohol: dependence syndr | 66590003 |
|  | E230.00 | Acute alcoholic intoxication in alcoholism | 191802004 |
|  | E231100 | Continuous chronic alcoholism | 191811004 |
|  | E011000 | Korsakov alcoholic psychosis | 69482004 |
|  | 1369.00 | Denies alcohol abuse | 413968004 |
|  | 8BA8.00 | Alcohol detoxification | 64297001 |
|  | Eu10611 | Korsakoff's psychosis | 69482004 |
|  | Eu10212 | [X]Chronic alcoholism | 66590003 |
|  | E231z00 | Chronic alcoholism NOS | 66590003 |
|  | E012.11 | Alcoholic dementia | 281004 |
|  | E231300 | Chronic alcoholism in remission | 191813001 |
|  | E250z00 | Nondependent alcohol abuse NOS | 268645007 |
|  | F375.00 | Alcohol-induced polyneuropathy | 7916009 |
|  | E231200 | Episodic chronic alcoholism | 191812006 |
|  | E010.00 | Alcohol withdrawal delirium | 8635005 |
|  | E01..00 | Alcohol-induced psychosis | 42344001 |
|  | E250200 | Nondependent alcohol abuse, episodic | 191883007 |
|  | J613000 | Alcoholic hepatic failure | 235881000 |
|  | E011200 | Wernicke-Korsakov syndrome | 69482004 |
|  | E01y.00 | Other alcoholic psychosis | 42344001 |
|  | 2577.11 | O/E - alcoholic breath | 163184002 |
|  | ZV11300 | [V]Personal history of alcoholism | 371422002 |
|  | E250100 | Nondependent alcohol abuse, continuous | 191882002 |
|  | F11x011 | Alcoholic encephalopathy | 192811002 |
|  | Eu10711 | [X]Alcoholic dementia NOS | 281004 |
|  | 13Y8.00 | Alcoholics anonymous | 1099951000000107 |
|  | F25B.00 | Alcohol-induced epilepsy | 361268000 |
|  | J612000 | Alcoholic fibrosis and sclerosis of liver | 235880004 |
|  | G852300 | Oesophageal varices in alcoholic cirrhosis of the liver | 309783001 |
|  | F144000 | Cerebellar ataxia due to alcoholism | 361272001 |
|  | Eu10411 | [X]Delirium tremens, alcohol induced | 8635005 |
|  | E012.00 | Other alcoholic dementia | 281004 |
|  | Eu10511 | Alcoholic hallucinosis | 7052005 |
|  | E013.00 | Alcohol withdrawal hallucinosis | 191476005 |
|  | E012000 | Chronic alcoholic brain syndrome | 191475009 |
|  | Eu10514 | [X]Alcoholic psychosis NOS | 42344001 |
|  | 1B1c.00 | Alcohol induced hallucinations | 417633001 |
|  | F11x000 | Cerebral degeneration due to alcoholism | 192811002 |
|  | F394100 | Alcohol myopathy | 19303008 |
|  | Eu10500 | Alcohol-induced psychosis | 42344001 |
|  | Eu10z00 | [X]Ment & behav dis due use alcohol: unsp ment & behav dis | 91388009 |
|  | 8G32.00 | Aversion therapy - alcoholism | 183388004 |
|  | J617000 | Chronic alcoholic hepatitis | 307757001 |
|  | E250300 | Nondependent alcohol abuse in remission | 191884001 |
|  | E230000 | Acute alcoholic intoxication, unspecified, in alcoholism | 191802004 |
|  | 7P22100 | Delivery of rehabilitation for alcohol addiction | 231161000000109 |
|  | E01z.00 | Alcoholic psychosis NOS | 42344001 |
|  | Eu10300 | [X]Mental and behav dis due to use alcohol: withdrawal state | 191480000 |
|  | E230z00 | Acute alcoholic intoxication in alcoholism NOS | 191802004 |
|  | E014.00 | Pathological alcohol intoxication | 191477001 |
|  | Eu10600 | Alcohol amnestic disorder | 73097000 |
|  | E011.00 | Alcohol amnestic syndrome | 69482004 |
|  | E015.00 | Alcoholic paranoia | 191478006 |
|  | E230200 | Episodic acute alcoholic intoxication in alcoholism | 191805002 |
|  | E230.11 | Alcohol dependence with acute alcoholic intoxication | 191802004 |
|  | Eu10y00 | [X]Men & behav dis due to use alcohol: oth men & behav dis | 29212009 |
|  | Eu10712 | [X]Chronic alcoholic brain syndrome | 191475009 |
|  | E011100 | Korsakov's alcoholic psychosis with peripheral neuritis | 191471000 |
|  | Eu10400 | [X]Men & behav dis due alcohl: withdrawl state with delirium | 8635005 |
|  | E230300 | Acute alcoholic intoxication in remission, in alcoholism | 191806001 |
|  | E230100 | Continuous acute alcoholic intoxication in alcoholism | 191804003 |
|  | Eu10700 | [X]Men & behav dis due alcoh: resid & late-onset psychot dis | 42344001 |
|  | C150500 | Alcohol-induced pseudo-Cushing's syndrome | 237738005 |
|  | Eu10513 | [X]Alcoholic paranoia | 42344001 |
|  | E011z00 | Alcohol amnestic syndrome NOS | 73097000 |
|  | Eu10512 | [X]Alcoholic jealousy | 42344001 |
|  | 8W2..00 | Referral to mental health services deferred until alcohol misuse resolved | 933641000000103 |

**(B)**

|  | **Read Code** | **Term** | **SNOMED CT Code** |
| --- | --- | --- | --- |
| **White** | 9i0..00 | British or mixed British - ethnic category 2001 census | 92391000000108 |
|  | 9i00.00 | White British - ethnic category 2001 census | 494131000000105 |
|  | 9i1..00 | Irish - ethnic category 2001 census | 92401000000106 |
|  | 9i2..00 | Other White background - ethnic category 2001 census | 92411000000108 |
|  | 9i10.00 | White Irish - ethnic category 2001 census | 494161000000100 |
|  | 9i20.00 | English - ethnic category 2001 census | 110761000000106 |
|  | 9i21.00 | Scottish - ethnic category 2001 census | 92541000000108 |
|  | 9i22.00 | Welsh - ethnic category 2001 census | 92551000000106 |
|  | 9i26.00 | Cypriot (part not stated) - ethnic category 2001 census | 92791000000109 |
|  | 9i27.00 | Greek - ethnic category 2001 census | 93931000000104 |
|  | 9i28.00 | Greek Cypriot - ethnic category 2001 census | 93941000000108 |
|  | 9i29.00 | Turkish - ethnic category 2001 census | 110401000000103 |
|  | 9i2A.00 | Turkish Cypriot - ethnic category 2001 census | 93951000000106 |
|  | 9i2B.00 | Italian - ethnic category 2001 census | 93961000000109 |
|  | 9i2C.00 | Irish Traveller - ethnic category 2001 census | 88911000000101 |
|  | 9i2D.00 | Traveller - ethnic category 2001 census | 88921000000107 |
|  | 9i2E.00 | Gypsy/Romany - ethnic category 2001 census | 88931000000109 |
|  | 9i2F.00 | Polish - ethnic category 2001 census | 88941000000100 |
|  | 9i2G.00 | Baltic States (Estonian or Latvian or Lithuanian) - ethnic category 2001 census | 88951000000102 |
|  | 9i2H.00 | Commonwealth of (Russian) Independent States - ethnic category 2001 census | 88961000000104 |
|  | 9i2J.00 | Kosovan - ethnic category 2001 census | 93981000000100 |
|  | 9i2K.00 | Albanian - ethnic category 2001 census | 88971000000106 |
|  | 9i2L.00 | Bosnian - ethnic category 2001 census | 93991000000103 |
|  | 9i2M.00 | Croatian - ethnic category 2001 census | 94001000000108 |
|  | 9i2N.00 | Serbian - ethnic category 2001 census | 88981000000108 |
|  | 9i2P.00 | Other republics which made up the former Yugoslavia - ethnic category 2001 census | 94011000000105 |
|  | 9i2Q.00 | Mixed Irish and other White - ethnic category 2001 census | 94021000000104 |
|  | 9i2R.00 | Other White European or European unspecified or Mixed European - ethnic category 2001 census | 94041000000106 |
|  | 9i24.00 | Northern Irish - ethnic category 2001 census | 92561000000109 |
|  | 9i25.00 | Ulster Scots - ethnic category 2001 census | 93921000000101 |
|  | 9i23.00 | Cornish - ethnic category 2001 census | 92571000000102 |
|  | 9i2S.00 | Other mixed White - ethnic category 2001 census | 94031000000102 |
|  | 9i2T.00 | Other White or White unspecified - ethnic category 2001 census | 94051000000109 |
|  | 9S1..00 | White | 185984009 |
|  | 9S10.00 | White British | 315236000 |
|  | 9S11.00 | White Irish | 315237009 |
|  | 9S12.00 | White - ethnic group | 185984009 |
|  | 9S13.00 | White Scottish | 401213008 |
|  | 9S14.00 | Other white British ethnic group | 401214002 |
|  | 9SA9.00 | Irish (NMO) | 186014006 |
|  | 9SAA.00 | Greek/Greek Cypriot (NMO) | 270466006 |
|  | 9SAA.11 | Greek (NMO) | 275599007 |
|  | 9SAA.12 | Greek Cypriot (NMO) | 275600005 |
|  | 9SAB.00 | Turkish/Turkish Cypriot (NMO) | 270467002 |
|  | 9SAB.11 | Turkish (NMO) | 275601009 |
|  | 9SAB.12 | Turkish Cypriot (NMO) | 275602002 |
|  | 9SI..00 | Irish traveller | 315283003 |
| **South Asians** | 9i7..00 | Indian or British Indian - ethnic category 2001 census | 110751000000108 |
|  | 9i8..00 | Pakistani or British Pakistani - ethnic category 2001 census | 92461000000105 |
|  | 9i9..00 | Bangladeshi or British Bangladeshi - ethnic category 2001 census | 92471000000103 |
|  | 9iA1.00 | Punjabi - ethnic category 2001 census | 92641000000107 |
|  | 9iA2.00 | Kashmiri - ethnic category 2001 census | 92651000000105 |
|  | 9iA4.00 | Sri Lankan - ethnic category 2001 census | 86461000000107 |
|  | 9iA5.00 | Tamil - ethnic category 2001 census | 92671000000101 |
|  | 9iA6.00 | Sinhalese - ethnic category 2001 census | 110781000000102 |
|  | 9S6..00 | Indian | 414481008 |
|  | 9S7..00 | Pakistani | 186002003 |
|  | 9S8..00 | Bangladeshi | 186003008 |
|  | 9SA7.00 | Indian sub-continent (NMO) | 186012005 |
| **Other Asians** | 9iA3.00 | East African Asian - ethnic category 2001 census | 92661000000108 |
|  | 9iA..00 | Other Asian background - ethnic category 2001 census | 92481000000101 |
|  | 9i64.00 | Asian and Chinese - ethnic category 2001 census | 92611000000106 |
|  | 9iA7.00 | Caribbean Asian - ethnic category 2001 census | 92691000000102 |
|  | 9iA8.00 | British Asian - ethnic category 2001 census | 92681000000104 |
|  | 9iA9.00 | Mixed Asian - ethnic category 2001 census | 92631000000103 |
|  | 9iAA.00 | Other Asian or Asian unspecified - ethnic category 2001 census | 92701000000102 |
|  | 9iF..00 | Other - ethnic category 2001 census | 92521000000101 |
|  | 9iE..00 | Chinese - ethnic category 2001 census | 92511000000107 |
|  | 9iF0.00 | Vietnamese - ethnic category 2001 census | 92751000000101 |
|  | 9iF1.00 | Japanese - ethnic category 2001 census | 92761000000103 |
|  | 9iF2.00 | Filipino - ethnic category 2001 census | 92771000000105 |
|  | 9iF3.00 | Malaysian - ethnic category 2001 census | 92781000000107 |
|  | 9iF9.00 | Arab - ethnic category 2001 census | 89001000000105 |
|  | 9iFB.00 | Middle Eastern (excluding Israeli, Iranian and Arab) - ethnic category 2001 census | 94071000000100 |
|  | 9iFD.00 | Iranian - ethnic category 2001 census | 89011000000107 |
|  | 9iFE.00 | Kurdish - ethnic category 2001 census | 94091000000101 |
|  | 9S9..00 | Chinese | 33897005 |
|  | 9SA4.12 | Iranian (NMO) | 275595001 |
|  | 9SA6.00 | E Afric Asian/Indo-Carib (NMO) | 270465005 |
|  | 9SA6.11 | East African Asian (NMO) | 275596000 |
|  | 9SA8.00 | Other Asian (NMO) | 186013000 |
|  | 9SC..00 | Vietnamese | 312859007 |
|  | 9SH..00 | Other Asian ethnic group | 315281001 |
| **Black** | 9iD3.00 | Mixed Black - ethnic category 2001 census | 92721000000106 |
|  | 9iD4.00 | Other Black or Black unspecified - ethnic category 2001 census | 92741000000104 |
|  | 9iB..00 | Caribbean - ethnic category 2001 census | 107691000000105 |
|  | 9iC..00 | African - ethnic category 2001 census | 92491000000104 |
|  | 9iD..00 | Other Black background - ethnic category 2001 census | 92501000000105 |
|  | 9iD0.00 | Somali - ethnic category 2001 census | 92711000000100 |
|  | 9iD1.00 | Nigerian - ethnic category 2001 census | 92731000000108 |
|  | 9iD2.00 | Black British - ethnic category 2001 census | 110791000000100 |
|  | 9iFH.00 | South and Central American - ethnic category 2001 census | 89021000000101 |
|  | 9S2..00 | Black Caribbean | 185988007 |
|  | 9S4..00 | Black, other, non-mixed origin | 185989004 |
|  | 9S3..00 | Black African | 18167009 |
|  | 9S41.00 | Black British | 185990008 |
|  | 9S42.00 | Black Caribbean/W.I./Guyana | 270460000 |
|  | 9S42.11 | Black Caribbean | 185988007 |
|  | 9S42.12 | Black West Indian | 309643000 |
|  | 9S42.13 | Black Guyana | 309644006 |
|  | 9S43.00 | Black N African/Arab/Iranian | 270461001 |
|  | 9S43.11 | Black North African | 275586009 |
|  | 9S43.12 | Black Arab | 275587000 |
|  | 9S43.13 | Black Iranian | 275588005 |
|  | 9S44.00 | Black - other African country | 185993005 |
|  | 9S45.00 | Black East African Asian/Indo-Caribbean | 270462008 |
|  | 9S45.11 | Black East African Asian | 275589002 |
|  | 9S45.12 | Black Indo-Caribbean | 275590006 |
|  | 9S47.00 | Black - other Asian | 185996002 |
|  | 9S48.00 | Black Black - other | 185989004 |
|  | 9S5..00 | Black - other, mixed | 185998001 |
|  | 9SA3.00 | Caribbean I./W.I./Guyana (NMO) | 270463003 |
|  | 9SA3.11 | Caribbean Island (NMO) | 275591005 |
|  | 9SA3.13 | Guyana (NMO) | 275593008 |
|  | 9SA3.12 | West Indian (NMO) | 275592003 |
|  | 9SG..00 | Other black ethnic group | 315279003 |
| **Mixed / Others** | 9SA4.00 | N African Arab/Iranian (NMO) | 270464009 |
|  | 9SA4.11 | North African Arab (NMO) | 275594002 |
|  | 9SA5.00 | Other African countries (NMO) | 186010002 |
|  | 9SA6.12 | Indo-Caribbean (NMO) | 275597009 |
|  | 9SAC.00 | Other European (NMO) | 186017004 |
|  | 9SAD.00 | Other ethnic NEC (NMO) | 186005001 |
|  | 9SB..00 | Other ethnic, mixed origin | 186019001 |
|  | 9SB1.00 | Other ethnic, Black/White orig | 186020007 |
|  | 9SB2.00 | Other ethnic, Asian/White orig | 186021006 |
|  | 9SB3.00 | Other ethnic, mixed white orig | 186022004 |
|  | 9SB4.00 | Other ethnic, other mixed orig | 186023009 |
|  | 9SB5.00 | Black Caribbean and White | 315634007 |
|  | 9SB6.00 | Black African and White | 315635008 |
|  | 9i65.00 | Other Mixed or Mixed unspecified - ethnic category 2001 census | 92621000000100 |
|  | 9i3..00 | White and Black Caribbean - ethnic category 2001 census | 92421000000102 |
|  | 9i4..00 | White and Black African - ethnic category 2001 census | 92431000000100 |
|  | 9i5..00 | White and Asian - ethnic category 2001 census | 92441000000109 |
|  | 9i6..00 | Other Mixed background - ethnic category 2001 census | 92451000000107 |
|  | 9i60.00 | Black and Asian - ethnic category 2001 census | 92581000000100 |
|  | 9i61.00 | Black and Chinese - ethnic category 2001 census | 92591000000103 |
|  | 9i62.00 | Black and White - ethnic category 2001 census | 110771000000104 |
|  | 9i63.00 | Chinese and White - ethnic category 2001 census | 92601000000109 |
|  | 9iFF.00 | Moroccan - ethnic category 2001 census | 94101000000109 |
|  | 9iFG.00 | Latin American - ethnic category 2001 census | 94111000000106 |
|  | 9iFJ.00 | Multi-ethnic islands: Mauritian or Seychellois or Maldivian or St Helena - ethnic category 2001 census | 94121000000100 |
|  | 9iFK.00 | Any other group - ethnic category 2001 census | 94151000000105 |
|  | 9S51.00 | Other Black - Black/White orig | 185999009 |
|  | 9S52.00 | Other Black - Black/Asian orig | 186000006 |

**(C)**

| **Variable Name** | **Read Code** | **Variable Name** | **Read Code** |
| --- | --- | --- | --- |
| **BMI** | **22K..00** | **GGT** | **44G9.00** |
| **ALT** | **44GB.00** |  | **44G4.00** |
| **AST** | **44HB.00** | **Cholesterol** | **44P..00** |
|  | **44H5.11** |  | **44PJ.00** |
|  | **44H5.00** | **LDL** | **44PI.00** |
| **HbA1c** | **42W5.00** |  | **44PE.00** |
|  | **44TB.00** | **Platelet count** | **42P..00** |
|  | **44TC.00** |  | **42PZ.00** |
|  | **42W..11** | **Triglycerides** | **44Q..00** |
|  | **42W4.00** |  | **44Q5.00** |
| **QRISK2** | **38DP.00** |  |  |

- **Reference:**
  - Kontopantelis E, Springate DA, Reeves D *et al.* *PH569 / 1138 - Type 2 Diabetes*. Phenotype Library [Online]. 06 October 2021. Available from: <http://phenotypes.healthdatagateway.org/phenotypes/PH569/version/1138/detail/>
  - Kuan V, Denaxas S, Gonzalez-Izquierdo A *et al.* *PH189 / 378 - Hypertension*. Phenotype Library [Online]. 06 October 2021. Available from: <http://phenotypes.healthdatagateway.org/phenotypes/PH189/version/378/detail/>
  - Carr MJ, Ashcroft DM, Kontopantelis E *et al. PH393 / 786 - Smoking*. Phenotype Library [Online]. 06 October 2021. Available from: <http://phenotypes.healthdatagateway.org/phenotypes/PH393/version/786/detail/>
  - Abel KM, Hope H, Swift E *et al*. *PH821 / 1721 - Alcohol Abuse*. Phenotype Library [Online]. 04 April 2022. Available from: <http://phenotypes.healthdatagateway.org/phenotypes/PH821/version/1721/detail/>
  - Reeves D, Springate DA, Ashcroft DM *et al.* *PH615 / 1230 - Body Mass Index*. Phenotype Library [Online]. 06 October 2021. Available from: <http://phenotypes.healthdatagateway.org/phenotypes/PH615/version/1230/detail/>
  - Fairhurst C, Watt I, Martin F *et al. PH448 / 896 - BMI*. Phenotype Library [Online]. 06 October 2021. Available from: <http://phenotypes.healthdatagateway.org/phenotypes/PH448/version/896/detail/>
  - Wright AK, Kontopantelis E, Emsley R *et al.* *PH714 / 1428 - Ethnicity*. Phenotype Library [Online]. 06 October 2021. Available from: <http://phenotypes.healthdatagateway.org/phenotypes/PH714/version/1428/detail/>
  - Rodgers LL, Weedon MN, Henley WE *et al.* *PH915 / 1909 - Type 2 Diabetes (Hba1C Test Codes)*. Phenotype Library [Online]. 04 April 2022. Available from: <http://phenotypes.healthdatagateway.org/phenotypes/PH915/version/1909/detail/>
  - Kontopantelis E, Springate D, Reeves D *et al.* *PH529 / 1058 - Hba1C Testing*. Phenotype Library [Online]. 06 October 2021. Available from: <http://phenotypes.healthdatagateway.org/phenotypes/PH529/version/1058/detail/>

**Supplementary Table 3. Characteristics of MASLD vs MASLD excluding patients with any ‘excess alcohol use’ record**

|  | MASLD (N = 365,797) | MASLD exclude patients with any excess alcohol use record (N=347,703) |
| --- | --- | --- |
| Age (years) | 53 (43 – 63) | 53 (43 - 63) |
| Sex – male | 51.8% | 50.9% |
| BMI (in kg/m^2^) | 31.3 (27.9 – 35.6) | 31.4 (28. 0 – 35.7) |
| Ethnicity |  |  |
| White | 215,220 (77.9%) | 202,064 (77.2%) |
| Black | 11,469 (4.2%) | 11,118 (4.2%) |
| South Asian | 30,505 (11.0%) | 30,031 (11.5%) |
| Other Asian | 14,078 (5.1%) | 13,860 (5.3%) |
| Mixed / Others | 4,928 (1.7%) | 4,771 (1.8%) |
| N/A | 89,597 (-) | 85,859 (-) |
| History of comorbidities |  |  |
| Had T2DM | 76,640 (21.0%) | 73,576 (21.2%) |
| Had hypertension | 129,156 (35.3%) | 122,431 (35.2%) |
| Smoked | 198,979 (54.4%) | 184,967 (53.2%) |
| Had excess alcohol use | 17,493 (4.8%) | - |
| Lab measurements |  |  |
| ALT (IU/L) | 38 (24 – 62) | 38 (24 – 62) |
| AST (IU/L) | 32 (24 – 45) | 31 (23 – 44) |
| Platelets (10^9^/L) | 255 (214 – 302) | 256 (215 - 302) |
| GGT (IU/L) | 59 (34 – 115) | 57 (33 – 108) |
| HbA1c (%) | 5.6 (4.5 – 6.2) | 38 (25 – 44) |
| Cholesterol (mmol/L) | 5.0 (4.2 – 5.9) | 5.0 (4.2 – 5.8) |
| Triglycerides (mmol/L) | 1.8 (1.3 – 2.5) | 1.8 (1.3 – 2.5) |
| LDL (mmol/L) | 2.9 (2.2 – 3.6) | 2.9 (2.2 – 3.6) |
| **Fibrosis-4 score** | 61,579 (16.8%) | 58,317 (16.8%) |
| Low risk (<1.30) | 41,222 (66.9%) | 39,569 (67.9%) |
| Indeterminate risk | 16,359 (26.6%) | 15,319 (26.3%) |
| High risk (>2.67) | 3,998 (6.5%) | 3,429 (5.9%) |

**Supplementary Table 4. The availability of characteristics and lab measurements of MASLD patients and matched controls in the CPRD, and pre-2015 vs post-2015.**

|  | Availability among all MASLD individuals | Availability among all Ctrl individuals | Availability among MASLD, pre-2015 | Availability among MASLD, post-2015 |
| --- | --- | --- | --- | --- |
| Total individuals | 365,797 (100%) | 1,460,288 (100%) | 116,897 (32% of all MASLD) | 248,900 (68% of all MASLD) |
| BMI | 262,133 (71.7%) | 964,306 (66.0%) | 75,579 (64.7%) | 186,554 (75.0%) |
| Ethnicity | 276,200 (75.5%) | 883,702 (60.5%) | 65,345 (55.9%) | 210,855 (84.7%) |
| ALT | 249,605 (68.2%) | 624,245 (42.7%) | 40,136 (34.3%) | 209,469 (84.2%) |
| AST | 83,449 (22.8%) | 200,528 (13.7%) | 17,749 (15.2%) | 65,700 (26.4%) |
| Platelets | 309,253 (84.5%) | 854,233 (58.5%) | 84,847 (72.6%) | 224,406 (90.2%) |
| GGT | 161,633 (44.2%) | 373,463 (25.6%) | 51,494 (44.1%) | 110,139 (44.3%) |
| HbA1c | 207,292 (56.7%) | 517,404 (35.4%) | 33,614 (28.8%) | 173,678 (69.8%) |
| Cholesterol | 268,049 (73.3%) | 724,697 (49.6%) | 75,686 (64.7%) | 192,363 (77.3%) |
| Triglycerides | 223,099 (61.0%) | 647,292 (44.3%) | 67,183 (57.5%) | 155,916 (62.6%) |
| LDL | 36,687 (10.0%) | 128,279 (8.8%) | 9,641 (8.2%) | 27,046 (10.9%) |

**Supplementary Table 5. Descriptive characteristics of MASLD patients between White Caucasians and South Asians**. (A) All individuals. (B) Individuals with low risk Fib-4 (Fib-4 <1.30). For categorical variables, records were reported as counts (in percentages); for continuous variables, records were reported as medians (with IQRs).

**(A)**

| All individuals | White Caucasians  (N = 215,220) | South Asians  (N = 30,505) | Test of difference (p-value) |
| --- | --- | --- | --- |
| Age (years) | 55 (45 – 65) | 46 (37 – 56) | <0.001 |
| Sex – male | 50.7% | 53.1% | <0.001 |
| BMI (in kg/m^2^) | 31.9 (28.3 – 36.2) | 28.9 (26.1 – 32.5) | <0.001 |
| History of comorbidities |  |  |  |
| Had T2DM | 44,776 (20.8%) | 9,511 (31.2%) | <0.001 |
| Had hypertension | 81,291 (37.8%) | 9,043 (29.6%) | <0.001 |
| Smoked | 137,648 (64.0%) | 9,660 (31.7%) | <0.001 |
| Had excess alcohol use | 12,810 (6.0%) | 446 (1.5%) | <0.001 |
| Lab measurements |  |  |  |
| ALT (IU/L) | 38 (24 – 62) | 38 (23 – 61) | <0.001 |
| AST (IU/L) | 32 (23 – 45) | 31 (23 – 43) | <0.001 |
| Platelets (10^9^/L) | 254 (213 – 300) | 270 (229 – 319) | <0.001 |
| GGT (IU/L) | 61 (34 – 119) | 45 (28 – 80) | <0.001 |
| HbA1c (%) | 5.6 (4.9 – 6.2) | 5.8 (5.3 – 6.6) | <0.001 |
| Cholesterol (mmol/L) | 5.0 (4.2 – 5.9) | 4.8 (4.1 – 5.6) | <0.001 |
| Triglycerides (mmol/L) | 1.8 (1.3 – 2.6) | 1.8 (1.3 – 2.5) | <0.001 |
| LDL (mmol/L) | 2.9 (2.2 – 3.6) | 2.7 (2.0 – 3.3) | 0.045 |
| **Fibrosis-4 score** | 42,803 (19.9%) | 4,467 (14.6%) | <0.001 |
| Low risk (<1.30) | 27,688 (64.7%) | 3,634 (81.4%) | - |
| Indeterminate risk | 12,019 (28.1%) | 724 (16.2%) | - |
| High risk (>2.67) | 3,096 (7.2%) | 109 (2.4%) | - |
| **QRISK2 score** | 67,870 (31.5%) | 10,966 (35.9%) | <0.001 |
| Low risk (<10%) | 34,662 (51.1%) | 6,478 (59.1%) | - |
| Medium risk | 19,547 (28.8%) | 2,392 (21.8%) | - |
| High risk (>20%) | 13,661 (20.1%) | 2,096 (19.1%) | - |

**(B)**

| Individuals with low risk Fib-4 | White Caucasians  (N = 27,688) | South Asians  (N = 3,634) | Test of difference (p-value) |
| --- | --- | --- | --- |
| Age (years) | 51 (40 – 59) | 43 (36 – 51) | <0.001 |
| Sex – male | 49.4% | 57.0% | <0.001 |
| BMI (in kg/m^2^) | 32.3 (28.7 – 36.7) | 28.7 (26.0 – 32.2) | <0.001 |
| History of comorbidities |  |  |  |
| Had T2DM | 5,153 (18.6%) | 1,038 (28.6%) | <0.001 |
| Had hypertension | 8,664 (31.3%) | 901 (24.8%) | <0.001 |
| Smoked | 17,080 (61.7%) | 1,188 (32.7%) | <0.001 |
| Had excess alcohol use | 1,353 (4.9%) | 39 (1.1%) | <0.001 |
| Lab measurements |  |  |  |
| ALT (IU/L) | 44 (27 – 66) | 47 (28 – 68) | <0.001 |
| AST (IU/L) | 28 (22 – 37) | 29 (23 – 39) | <0.001 |
| Platelets (10^9^/L) | 274 (238 – 317) | 281 (243 – 327) | <0.001 |
| GGT (IU/L) | 52 (31 – 91) | 44 (29 – 73) | <0.001 |
| HbA1c (%) | 5.6 (5.3 – 6.2) | 5.8 (5.4 – 6.5) | <0.001 |
| Cholesterol (mmol/L) | 5.2 (4.4 – 6.0) | 4.9 (4.2 – 5.6) | <0.001 |
| Triglycerides (mmol/L) | 1.9 (1.4 – 2.7) | 1.8 (1.3 – 2.6) | <0.001 |
| LDL (mmol/L) | 3.1 (2.4 – 3.7) | 2.9 (2.2 – 3.5) | <0.001 |

**Supplementary Figure 1. The study flow chart.**

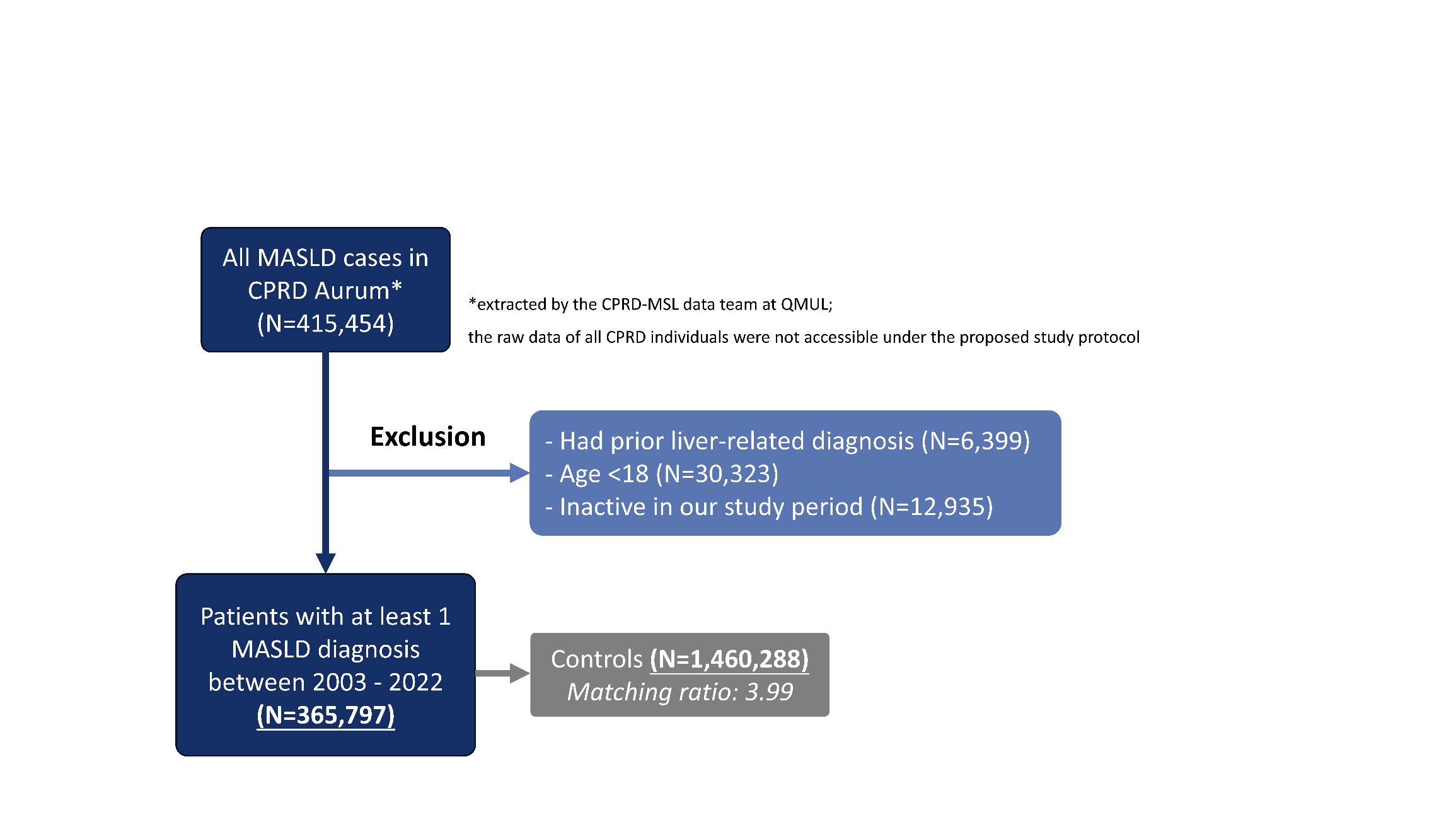

**Supplementary Figure 2. Age and sex subgroups of recorded incidence of MASLD in the CPRD, in 2007, 2012, 2017, and 2022.**

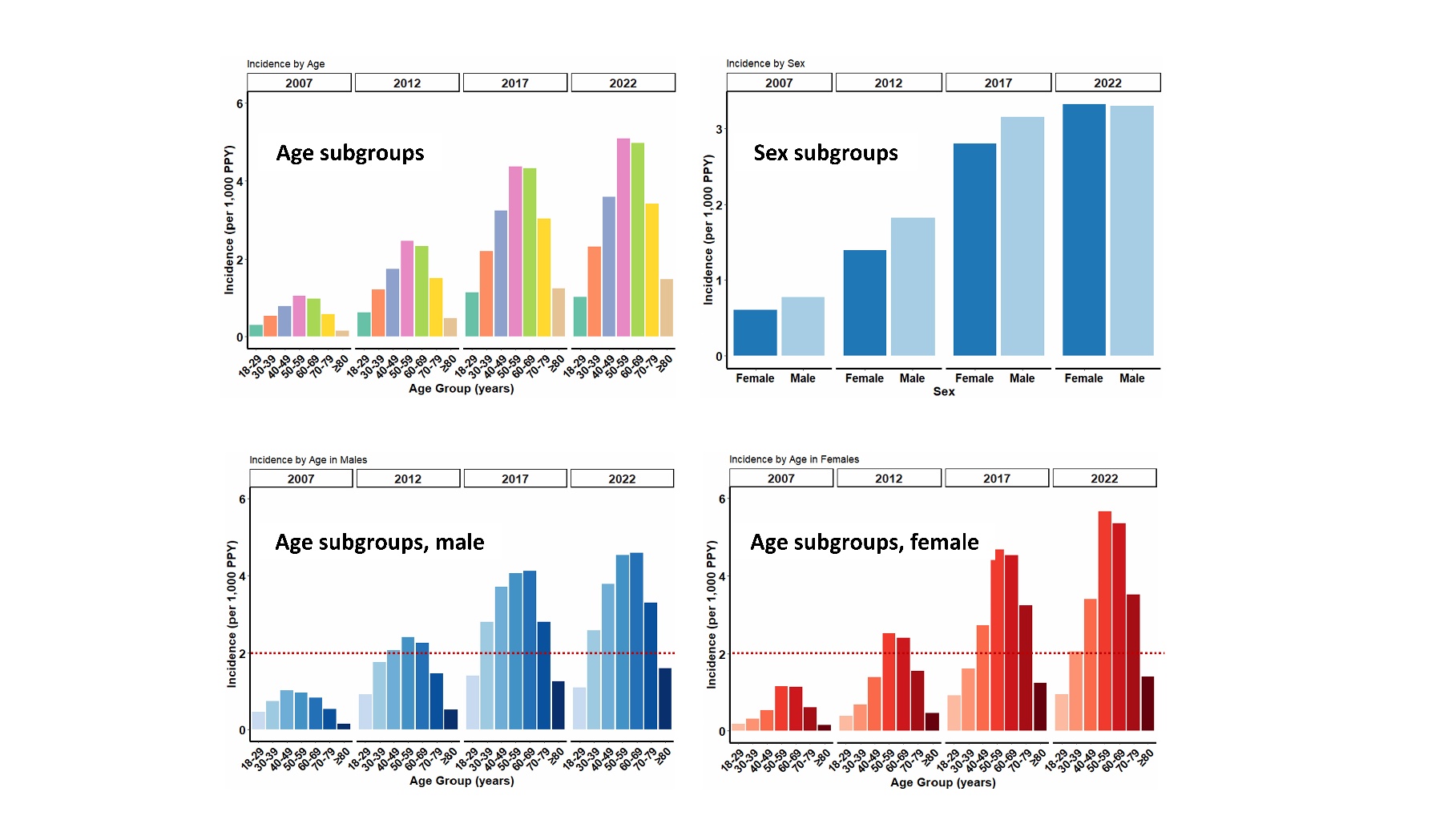

**Supplementary Figure 3. Annual incidence proportion of MASLD between 2003 and 2022, and age and sex subgroups in 2007, 2012, 2017, and 2022.**

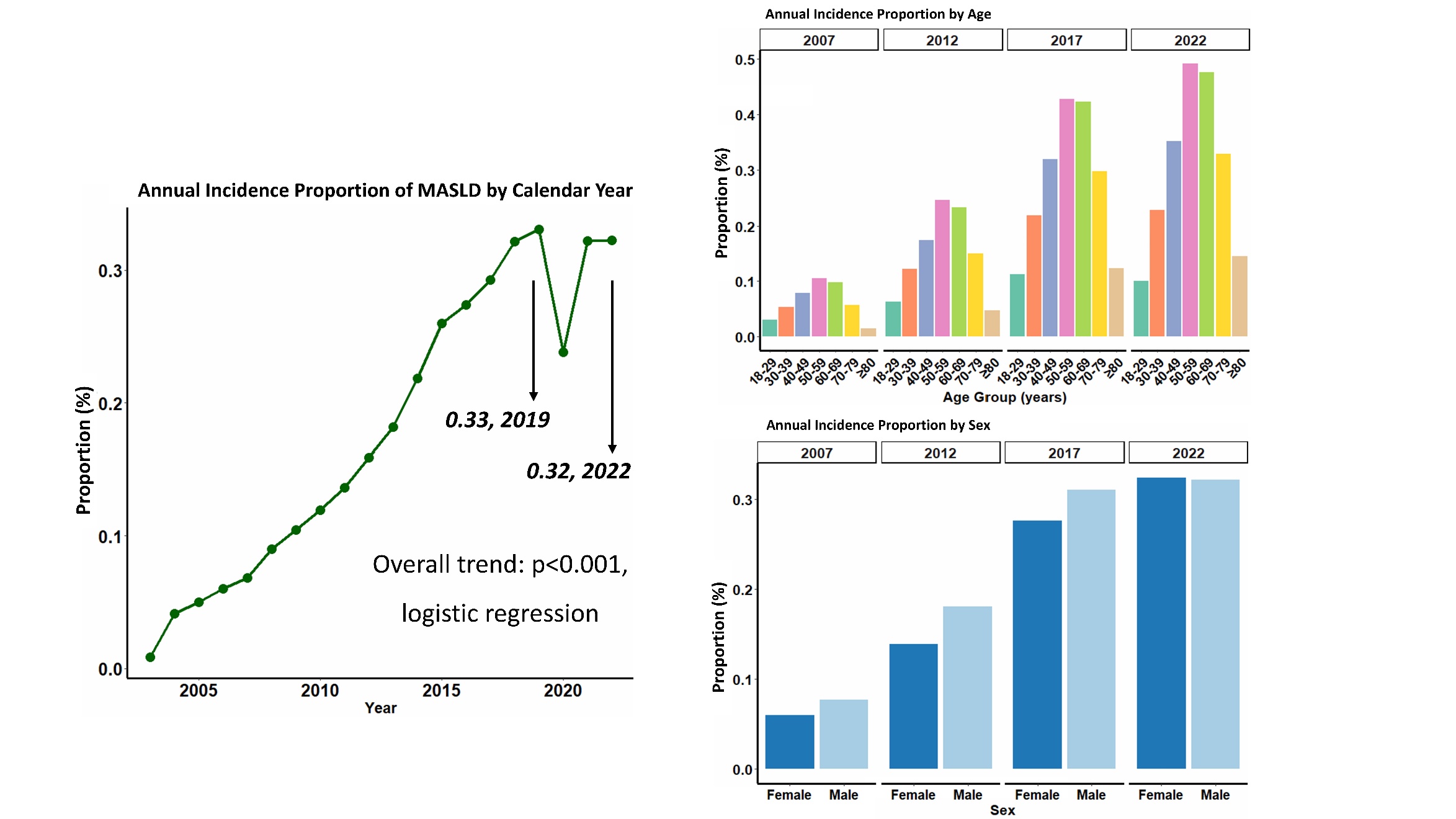

**Supplementary Figure 4. Distribution of time points for each Fib-4 component in MASLD patients.**

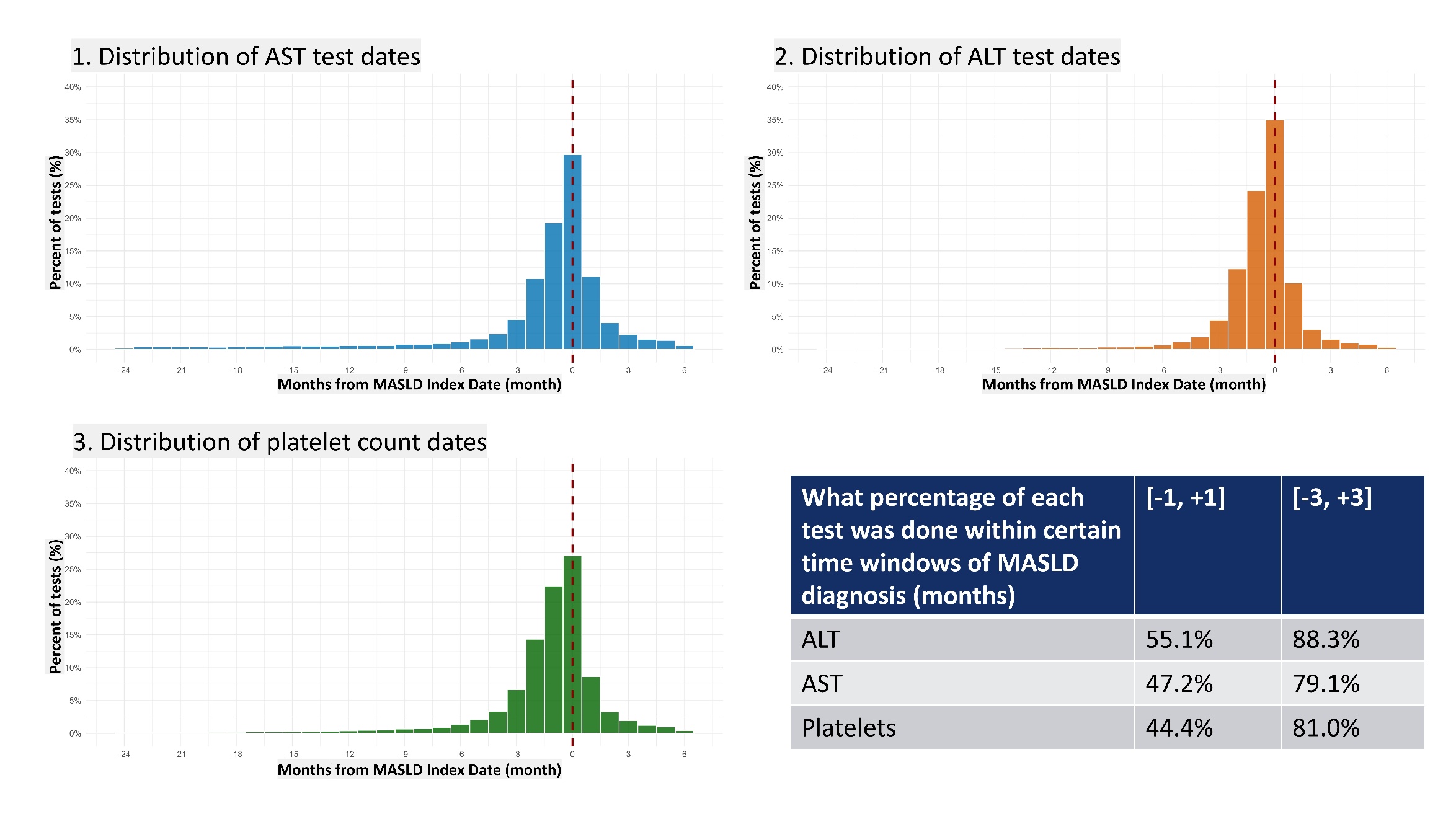

**Supplementary Figure 5. Temporal trend of Fib-4 components availability at the time of MASLD diagnosis, 2003 to 2022.** An effective test is defined as the closest record within [-2 years, +6 months] of the MASLD index date. (A) Overall trend. (B) Racial-ethnic trend.
**(A)**

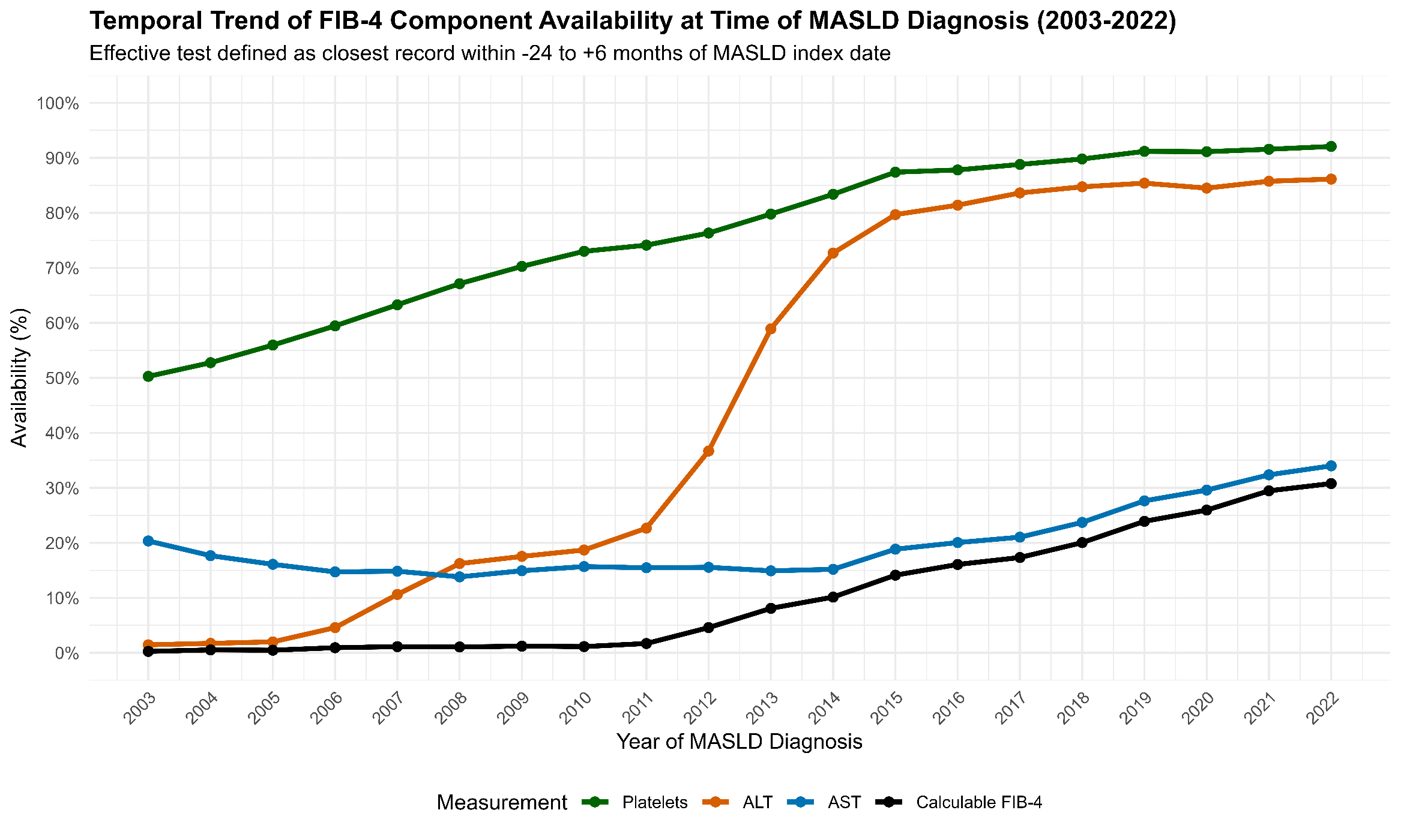

**(B)**

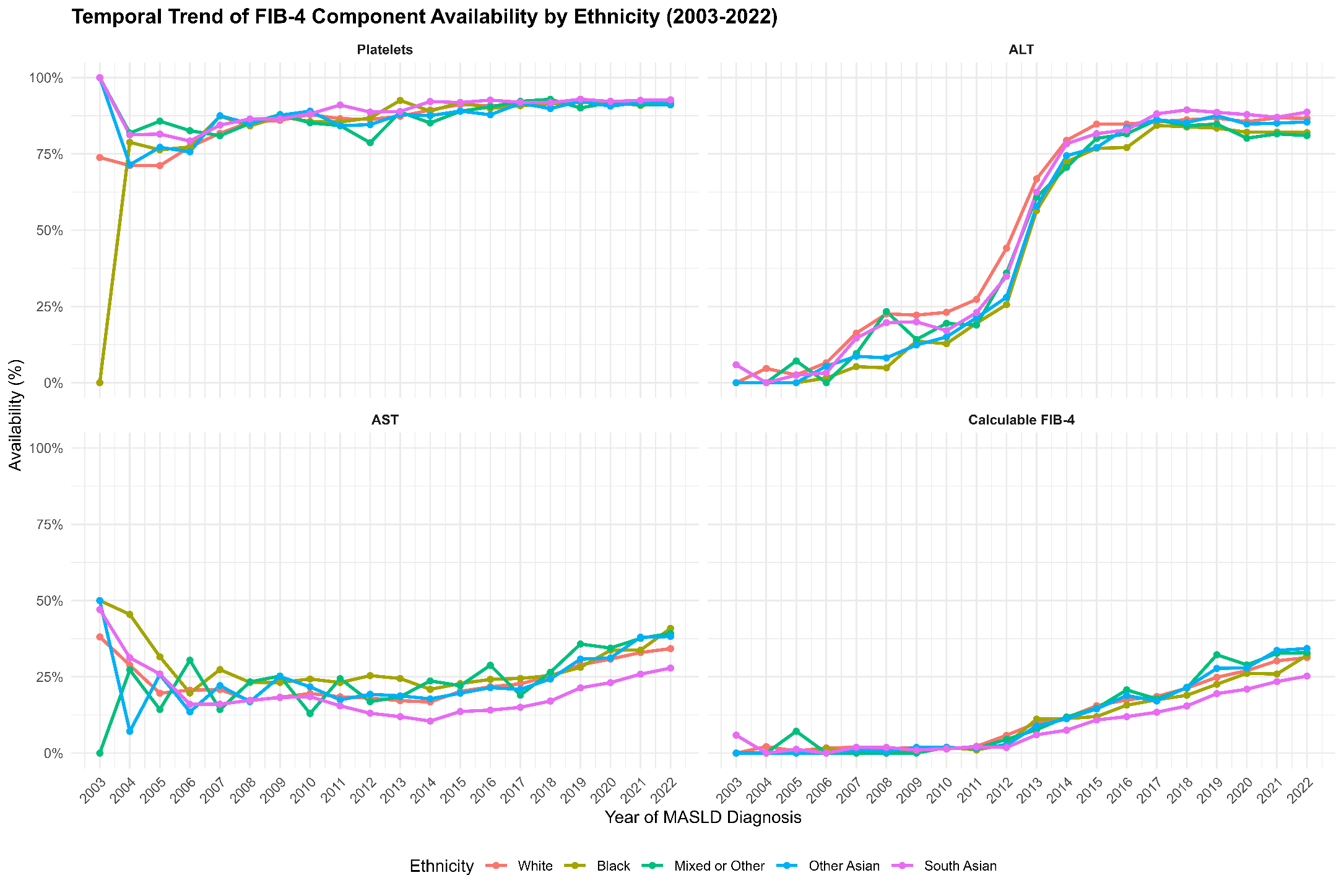
